# Mendelian gene identification through mouse embryo viability screening

**DOI:** 10.1101/2022.01.07.22268899

**Authors:** Pilar Cacheiro, Carl Henrik Westerberg, Jesse Mager, Mary E. Dickinson, Lauryl M.J. Nutter, Violeta Muñoz-Fuentes, Chih-Wei Hsu, Ignatia B. Van den Veyver, Ann M. Flenniken, Colin McKerlie, Stephen A. Murray, Lydia Teboul, Jason D. Heaney, K. C. Kent Lloyd, Louise Lanoue, Robert E. Braun, Jacqueline K. White, Amie K. Creighton, Valerie Laurin, Ruolin Guo, Dawei Qu, Sara Wells, James Cleak, Rosie Bunton-Stasyshyn, Michelle Stewart, Jackie Harrisson, Jeremy Mason, Hamed Haseli Mashhadi, Helen Parkinson, Ann-Marie Mallon, International Mouse Phenotyping Consortium, Genomics England Research Consortium, Damian Smedley

## Abstract

The diagnostic rate of Mendelian disorders in sequencing studies continues to increase, along with the pace of novel disease gene discovery. However, variant interpretation in novel genes not currently associated with disease is particularly challenging and strategies combining gene functional evidence with approaches that evaluate the phenotypic similarities between patients and model organisms have proven successful.

A full spectrum of intolerance to loss-of-function variation has been previously described, providing evidence that gene essentiality should not be considered as a simple and fixed binary property. Here we further dissected this spectrum by assessing the embryonic stage at which homozygous loss-of-function results in lethality in mice from the International Mouse Phenotyping Consortium, classifying the set of lethal genes into one of three windows of lethality: early, mid or late gestation lethal.

We studied the correlation between these windows of lethality and various gene features including expression across development, paralogy and constraint metrics together with human disease phenotypes, and found that the members of the early gestation lethal category show distinctive characteristics and a strong enrichment for genes linked with recessive forms of inherited metabolic disease.

Based on these findings, we explored a gene similarity approach for novel gene discovery focused on this subset of lethal genes. Finally, we investigated unsolved cases from the 100,000 Genomes Project recruited under this disease category to look for signs of enrichment of biallelic predicted pathogenic variants among early gestation lethal genes and highlight two novel candidates with phenotypic overlap between the patients and the mouse knockout.

## Introduction

The rate of molecular diagnosis through genomics approaches continues to improve. However, the diagnostic yield for Mendelian disorders varies significantly, ranging from 25 to 58%^1,2^ depending on the age of the proband, the type of disorder, the criteria for patient inclusion (e.g. absence of a clear clinical diagnosis, previous attempts to provide a molecular diagnosis) and the availability of sequence data from family members e.g. familial versus sporadic cases. Despite this progress, a considerable proportion of patients remain without a diagnosis. Potential strategies to address the challenge of undiagnosed patients and advance our understanding of the molecular basis of these disorders include but are not limited to: i) identifying novel Mendelian disease genes^3^, ii) developing experimental and computational approaches to assess the pathogenicity of variants of unknown significance in known disease genes, iii) considering expansion of the phenotype of known disease genes^4^, iv) investigating noncoding, regulatory variants, v) assessing the contribution of structural variation^5^, vi) investigating somatic mosaicism and, vii) exploring alternative modes of inheritance, i.e. digenic or multigenic^2^.

With regard to the first approach, the number of genes currently known to be associated with rare disorders comprises 20-25% of the protein coding genome according to OMIM^6^. There are between 200-300 new disease-gene associations published every year^7^, with many more to be uncovered. The number of additional disease-associated genes yet to be identified is estimated to be high, up to 1.5-3 times the number of currently known causative genes of Mendelian conditions^8^.

Combining different sources of information can boost the evidence for new associations. Integrating research and clinical datasets has proven to be effective at discovering the molecular basis for genetic disorders^9,10^. Model organism information on viability and cross-species phenotype comparisons in combination with clinical data constitutes another powerful strategy. Some examples include the automatic detection of mouse models for human disease and phenotype based variant prioritisation using algorithms such as PhenoDigm and Exomiser^11–13^. Additionally, mouse data on essentiality can be used as a discovery and prioritisation tool^14,15^. We previously developed a gene prioritisation strategy focused on neurodevelopmental disorders by integrating evidence of intolerance to loss-of-function (LoF) variation from multiple resources and bringing in data from large scale sequencing programs^16^. Through this approach combining viability data from mice and human cell line screens, we were able to identify a set of developmentally lethal genes, i.e. genes not essential for cell proliferation but required for organism development, which were enriched for autosomal dominant, developmental disease-associated genes. Investigation of clinical cases with *de novo* variants in developmental lethal genes and phenotypic overlap between the knockout mouse and affected individuals led us to prioritise a set of 9 candidate genes. Two of these genes have since been validated^17,18^.

To improve and expand these successful strategies to other types of disorders, here we again leverage evidence from high-throughput mouse phenotype screens conducted by the International Mouse Phenotyping Consortium (IMPC) to further explore the spectrum of intolerance to LoF variation. For genes with null alleles that result in a lethal phenotype in a primary viability screen (i.e. no live homozygous animals detected at weaning), the IMPC performs a secondary embryo viability screen to determine a ‘window of lethality’ (WoL). These WoL were defined by examining the survival of homozygous null mutants at up to four embryonic developmental time points: embryonic day (E) 9.5, E12.5, E15.5, and E18.5, with the WoL being the interval between the last stage at which homozygous null embryos are identified and the next latest examined time point^15^. In the present study we further dissected this set of lethal genes in the mouse with the primary aim of investigating how they can inform human disease gene discovery.

First, we explored these WoL and show how they relate to essentiality inferred from human cell proliferation assays, gene expression across development, various intolerance to variation metrics and duplication events. Secondly, we investigated these WoL in the context of human Mendelian disease and found that the set of early-gestation lethal genes in the mouse shows a strong correlation with autosomal recessive disease associated genes, in particular those involved in inherited metabolic disorders, resulting mainly from enzyme deficiencies^19^. Thirdly, we built a classifier to predict new early-gestation lethal genes and developed a strategy using gene similarity to biallelic inborn errors of metabolism genes (BIEM), a broad category of genes that function in metabolism and impact, or are impacted by most cellular processes^20^ and describe new candidate genes for these type of disorders. Finally, we explored unsolved metabolic disorder cases from the 100,000 Genomes Project (100KGP)^21^ to look for enrichment of biallelic predicted pathogenic variants among those genes, and provide a set of prioritised novel genes with shared phenotypes between patients and mouse knockouts.

## Results

### 1. Gaining functional knowledge from WoL

The IMPC measures viability at wean, and for lethal strains employs a high-throughput embryonic phenotyping pipeline to examine embryo viability and phenotypes at E9.5, E12.5, E15.5, and E18.5. The developmental period during which lethality occurs in the mouse can be summarised by establishing a set of WoL. A WoL for a gene was defined by the interval between the latest developmental stage at which live homozygous null embryos (mice) are identified and the earliest stage at which no live homozygous embryos are found. Complete lethality by E9.5 was classified as early-gestation lethal (EL), by E12.5 or E15.5 as mid-gestation lethal (ML), and viability at E15.5 or E18.5 as late-gestation lethal (LL). These WoL approximately correlate with the pre-organogenesis, organogenesis and post-organogenesis phases of mouse embryonic development, while also providing sufficient sample sizes to perform downstream statistical analyses. Among 895 embryonic lethal genes with one-to-one human orthologues, nearly half (430, 48%) are EL, 155 (17%) ML, and 310 (35%) are LL. A full description of the WoL and the distribution of lines per window can be found in **Fig 1** and **Sup File 1**.

**Fig 1.**
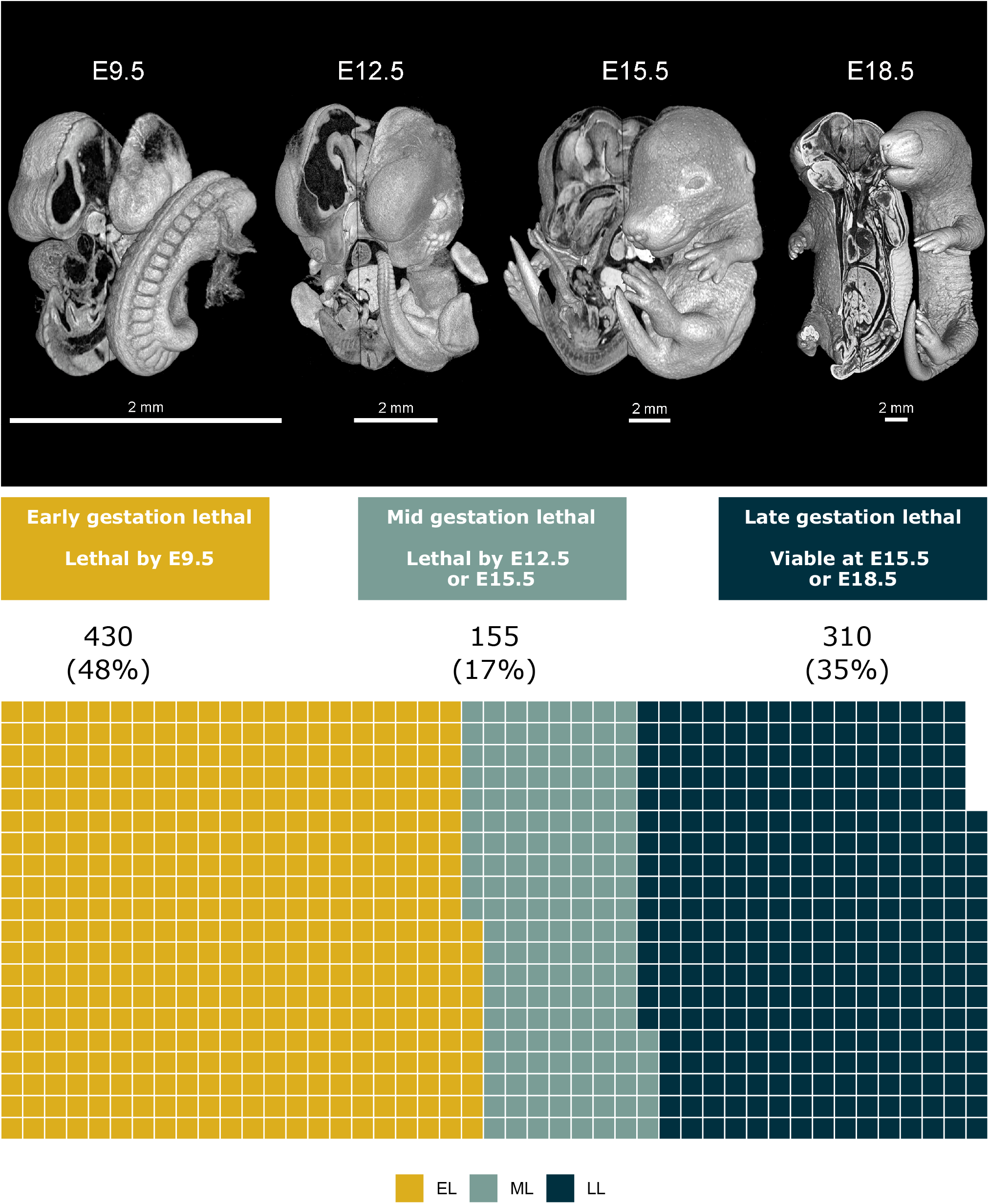
Description of the WoL and distribution of lethal genes across windows. Three dimensional microCT images of wild type mouse embryos corresponding to E9.5, E12.5, E15.5 and E18.5. The waffle chart shows the total number of lethal lines characterised through the secondary viability screening and their distribution by WoL. EL genes, where the embryo dies before embryonic day 9.5 constitute nearly 50% of all the lethal genes in the mouse. This stage broadly correlates with the pre-organogenesis phase of embryonic development. Non-early lethal lines are divided into ML (17%) and LL (35%). The complete set of genes associated with each WoL is available in **Sup File 1**. WoL, windows of lethality; EL, early gestation lethal; ML, mid gestation lethal; LL, late gestation lethal.

#### 1.1. Human cellular essential genes correlate with mouse EL genes

We previously reported that EL genes show a considerable overlap with human cellular essential genes^16^. Plotting individual gene-based proliferations scores for different human cell lines across tissues obtained from CRISPR knockout screens through the Achilles pipeline^22^, we observed a clear distinction between the three WoL. The set of EL genes stand alone as a distinctive category from the ML and LL genes that show closer median values (shown in **Fig 2a** for central nervous system cell lines, other lineages in **Sup Fig 1a-b**). Considering the average CERES score across cell lines, where lower values indicate more depletion and higher essentiality, we observed that all the genes are EL genes for the bins with lowest scores, and that the percentage of ML and LL genes increased as we move towards higher values of this score (**Fig 2b**). When cellular essentiality is considered as a binary property after categorising the mean scores using a cut-off of −0.45 (≤ −0.45: “cellular essential”, >-0.45: “cellular non-essential”, see Methods), 73% of EL genes are essential in human cell lines, compared to 25% of ML genes and only 6% of LL genes (**Fig 2c**). Alternative thresholds are considered in **Sup Fig 1c-d**.

**Fig 2.**
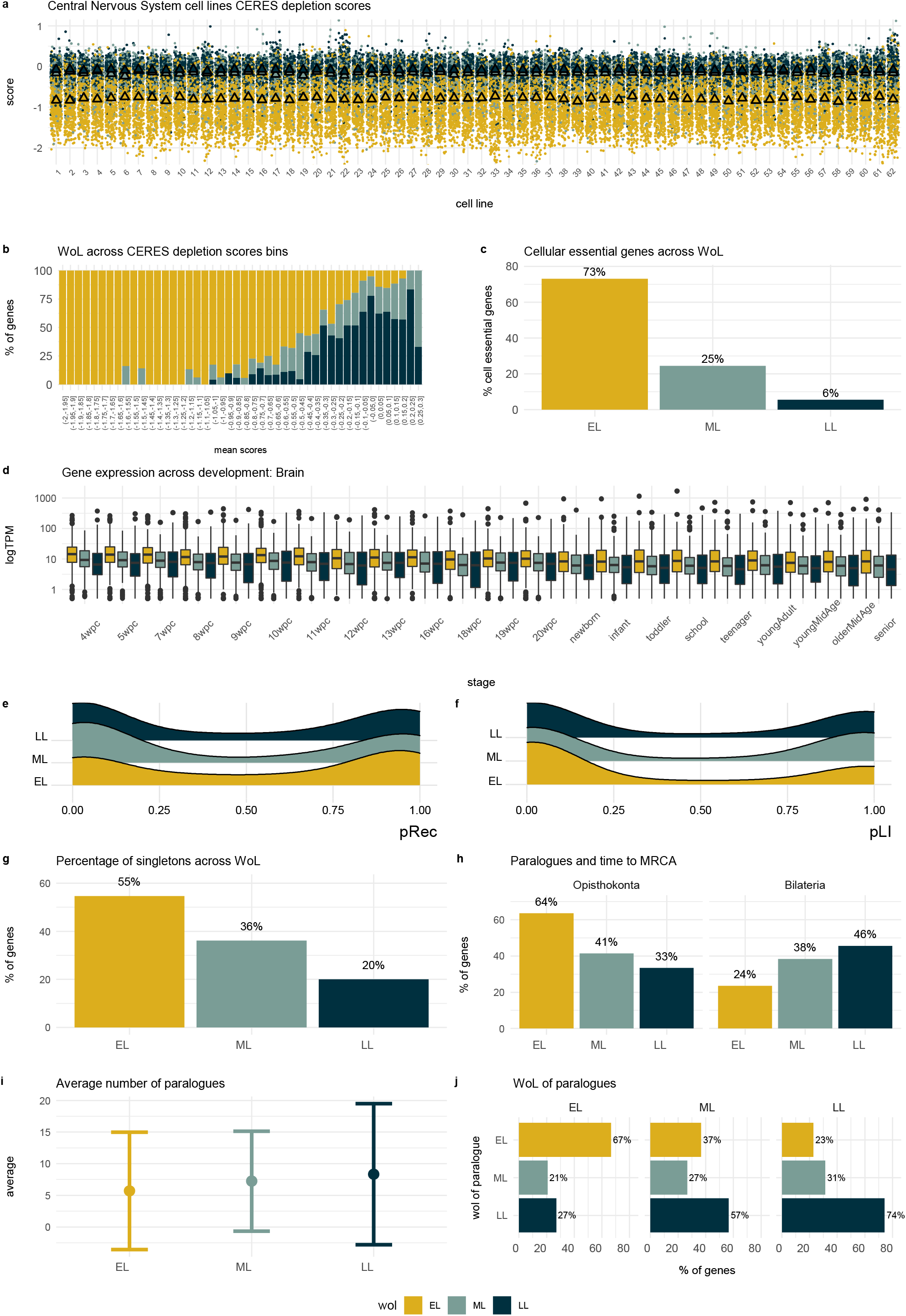
WoL and gene features. **2a CERES depletion scores for different Central Nervous System human cell lines across WoL.** A more negative scores indicates more depletion of the gene in the cell line, i.e. more essential. Triangles represent median values of gene expression per WoL. **2b Distribution of EL, ML and LL genes across mean CERES depletion scores bins**, with lower values covered 100% by EL genes. **2c WoL and cellular essential genes**. Percentage of EL, ML and LL genes considered cellular essential when a mean CERES depletion scores across cell lines of −0.45 is considered as threshold. **2d Brain gene expression**. Boxplots showing the distribution of human gene expression values across development stages for brain tissue. **2e Intolerance to homozygous LoF variation**. Distribution of the probability of being intolerant to homozygous LoF variation (pRec) scores across WoL, showing the bimodal distribution of this score and the underrepresentation of low pRec values among the EL. **2f Intolerance to heterozygous LoF variation**. Distribution of the probability of being LoF intolerant (pLI) scores across WoL, showing the bimodal distribution of this score and the underrepresentation of high pLI values among the EL. **2g WoL and singletons**. Percentage of singletons across WoL, with the proportion of genes with no duplicates decreasing across development stages. **2h Paralogues and time of the duplication event**. Paralogues of EL genes have an older origin, with a more ancient time to the most recent common ancestor. **2i Average number of paralogues**. Distribution of the average number of paralogues per gene for those genes with duplicates. **2j WoL of paralogues**. Paralogues of EL genes are more likely to be EL, paralogues of ML and LL are more likely to be LL. Tests for differences between windows available in **Sup Table 1**. For plots **2a – 2j**, the data shown correspond to gene annotations for the human orthologues. WoL, windows of lethality; EL, early gestation lethal; ML, mid gestation lethal; LL, late gestation lethal; MRCA, most recent common ancestor.

#### 1.2. EL genes consistently show higher levels of human gene expression across organs and developmental stages

Examination of human gene expression data^23^ showed a consistent pattern across organs and developmental stages with the human orthologues of mouse EL genes being expressed at higher levels, on average, compared to the orthologues of mouse ML and LL genes (**Fig 2d**). High levels of expression may help identify key developmental process. To that end, gene expression patterns during early human development have been used to predict essential genes lacking a known human disease association^24^. To assess whether the organ development trajectories for these genes differ substantially between mouse and human, we investigated the similarity of spatiotemporal gene expression profiles for the two species. We found that 78 and 82% of the entire set of genes under study showed the same trajectory for cerebellum and brain respectively, with no significant differences observed between WoL, and in concordance with what was observed for the entire set of genes with data available^25^ (**Sup Fig 2**). Similarities in gene expression will not always imply conserved phenotypes between mouse and human, but they can serve as a proxy for how translatable to human disease the findings for these genes are.

#### 1.3. Intolerance to LoF variation differs across WoL

EL genes are more intolerant to homozygous LoF variation based on human population sequencing data when compared to LL genes, as shown by a higher frequency of probability of intolerance to homozygous LoF variation (pRec) values close to 1 (**Fig 2e**). Consistently, human orthologues of EL genes also show an underrepresentation among genes with intolerance to heterozygous LoF as indicated by a lower frequency of high probability of intolerance to heterozygous (pLI) scores (**Fig 2f**). Albeit not statistically significant, this observation agrees with our previous findings that developmental lethal genes, which broadly correlate with ML or LL genes are more intolerant to heterozygous LoF variation. Similar results were obtained when we explored DOMINO scores that compute the likelihood of a gene to be associated with autosomal dominant disorders, i.e. EL genes were more likely to be linked to autosomal recessive disease compared to LL genes (**Sup Fig 3a**).

#### 1.4. Gene duplicates and time of duplication event are distinctive features of EL genes

EL genes have the highest proportion of genes with no paralogues (singletons). This proportion decreases gradually from ML to LL genes (**Fig 2g**). Not only are EL genes more likely to be singletons, but also, for those genes that do have paralogues, the number of paralogues is lower and the paralogues are more likely to be older, with longer times tracking back to the duplication event when compared to ML or LL genes, which suggests more time to evolve new functions (**Fig 2h, 2i**). Thus, not only do gene duplications, or the lack thereof, seem to play a role in essentiality but so do the number of paralogues and the time of the duplication event. Similar observations were made by others using different species and/or definitions of essentiality^26,27^. Paralogues of EL genes are also more likely to be EL, and similarly paralogues of ML/LL genes are more likely to be ML/LL. This implies that paralogues are predominantly essential at the same developmental stage, potentially reflecting similar key functions at the cellular level and early stages of organism development (**Fig 2j**). Additionally, when genes are divided into singletons and duplicates, the proportion of genes that are cellular essential is higher among the singletons compared to those genes with paralogues, and this observation is consistent for the three WoL (**Sup Fig 3b**). However, other studies investigating the relationship between essentiality, developmental expression, and gene duplication have suggested that timing of developmental expression influences the ability of a gene in a paralogue pair to compensate for the loss of function of the other gene^28^.

### 2. WoL and Mendelian disease

It is well established that there is an association between lethal genes in the mouse and human disease genes^15,29^. Our previous study showed that this enrichment was mainly driven by developmental lethal genes^16^ so we hypothesised that the distribution of disease genes across WoL may not be uniform and that information about WoL could highlight additional correlations. When translating our WoL to relevant developmental stages in humans, the EL mouse category broadly correlates with the human pre-organogenesis stage occurring during the first two weeks of development. The ML class relates to human organogenesis occurring during the embryonic period from weeks three through eight, and ending in the early first trimester, around week nine of gestation. Lastly, the LL category aligns with the human foetal stage, from the ninth week until birth^30^.

We used the Genomics England PanelApp, a publicly available knowledgebase containing expert curated gene panels related to human disorders, as the source of Mendelian genes to perform subsequent analyses^31^. Genes are rated according to level of evidence to support the phenotype association: ‘green’ means high level of evidence from several unrelated families and/or strong additional functional data, ‘amber’ moderate evidence, and ‘red’ not enough evidence. The advantages of using this source of diagnostic genes include the high level disease categorisation and allelic requirement annotations that allows for tailored analysis, the categorisation of genes according to the level of evidence for the gene-disease association, and the potential to map directly to patient data recruited in the 100KGP.

#### 2.1. Disease category and mode of inheritance are not uniformly distributed among WoL

Although the three WoL are all enriched for Mendelian disease genes, their properties differ. The proportion of genes associated with rare disorders is lowest among the EL, followed by the ML and LL genes (**Fig 3a**). When allelic requirement is considered, a similar pattern is found for monoallelic (dominant) disease-associated genes. However, this trend is reversed for recessive disorder-associated genes, where the EL fraction showed a significantly higher number of biallelic genes (**Fig 3b**).

**Fig 3.**
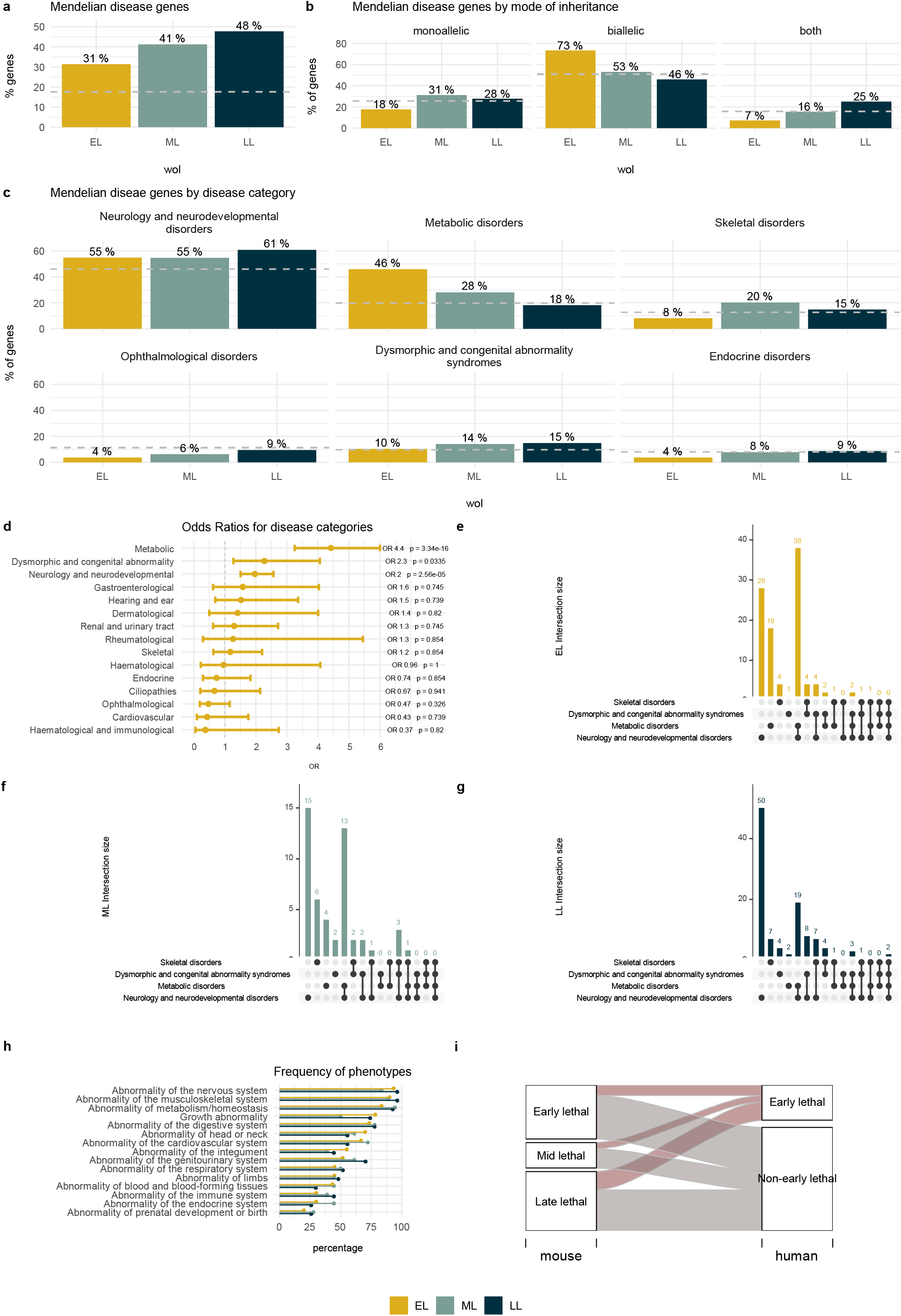
WoL and human disease. **3a Mendelian disease genes.** Percentage of rare disease associated genes in each WoL according to PanelApp, only ‘green’ genes with high level of evidence for the disease association are included. **3b Mode of inheritance**. Distribution of disease genes across WoL by associated allelic requirement, only monoallelic or biallelic patterns of inheritance are included. **3c Disease class**. Distribution of disease genes across WoL by disease type according to PanelApp level 2 disease categories, with the bars indicating the percentage of PanelApp genes mapping each disease class for the 3 WoL. For plots **3a, 3b** and **3c**, the dashed grey line represents the baseline percentage for the entire set of protein coding genes (19,197 genes according to HGNC, **3a**) or PanelApp ‘green’ genes (3,384 genes, **3b, 3c**). **3d Disease categories Odds Ratios** and BH adjusted P values for EL genes compared to ANEL genes: this includes mid and late gestation lethal genes as well as subviable and viable categories. **3e, 3f, 3g Disease category overlap**. Overlap between genes associated with the most frequent disease categories across WoL for EL, ML and LL genes respectively. **3h Top level HPO annotations**. Frequency of top-level HPO phenotype annotations for inborn errors of metabolism genes in each window. **3i WoL and early lethality in humans**. Human early lethal genes: PanelApp ‘green’ genes associated to early lethality (see Methods); human non early lethal genes: the remaining set of PanelApp ‘green’ genes. Tests for differences between WoL are available in **Sup Table 2**. WoL, windows of lethality; EL, early gestation lethal; ML, mid gestation lethal; LL, late gestation lethal; ANEL, all non-early gestation lethal genes; HPO, human phenotype ontology.

Further dissection of disease genes according to PanelApp high level disease categories showed that: 1) the proportion of neurodevelopmental disorder associated genes is higher than expected among the three WoL compared to baseline, with the highest percentage among LL genes, 2) the proportion of genes associated to metabolic disorders follows the inverse pattern, with EL genes showing the highest percentage of inherited metabolic disease genes (46%), followed by ML (28%) and showing the lowest percentage among the LL (18%) (most notably, this is the only disease category where we found a higher percentage of disease genes among the EL compared to ML and LL), 3) skeletal disorders are predominant among the ML, and 4) for the remaining disease categories, the frequency of disease genes among the EL genes show values comparable to baseline or even lower, indicative of depletion of these disease categories among the EL genes (**Fig 3c**). Only the 6 most frequent disease categories are shown here (additional disease categories in **Sup Fig 4**). Odds Ratio were computed for EL genes, where non-EL genes included ML, LL, subviable, and viable categories (See Methods). Three disease categories showed significant OR > 1: metabolic disorders (OR = 4.4; P value = 3.34e-16), dysmorphic and congenital abnormality syndromes (OR = 2.3; P value = 0.034), and neurology and neurodevelopmental disorders (OR = 2; P value = 2.56e-05) (**Fig 3d**).

Given that most inborn errors of metabolism (IEM) show neurological manifestations, and neurodevelopmental disorders are still the most predominant disease category across the three WoL, we further explored the gene overlap between neurodevelopmental and metabolic disease categories to assess any potential confounding effect. The combination of genes associated with both metabolic and neurodevelopmental disorders was found to be predominant among the EL class, opposite to what we observed among the ML and LL windows, where neurodevelopmental only genes are the prevalent disease class, and thus providing additional evidence for the inborn errors of metabolism association (**Fig 3e, 3f, 3g**).

The analysis of Human Phenotype Ontology (HPO) phenotypes associated with known inborn error of metabolism genes showed that the five most frequent physiological systems affected are: nervous system, followed by musculoskeletal, metabolism/homeostasis, growth abnormality, and digestive. An enrichment analysis showed no significant differences in the frequency of any particular phenotype for EL genes when compared to ML and LL (**Fig 3h, Sup Table 3**).

#### 2.2. Evidence of prenatal and perinatal lethality in humans

Among the wide range of Mendelian phenotypes observed in humans, prenatal lethality poses a unique challenge in terms of providing a molecular diagnosis. Development failure may occur at any point between fertilisation and birth. Estimates suggest that 20-30% of implanted embryos fail to develop beyond week six^32^, similarly early embryo losses occurring between implantation and clinical recognition could be around 10–25%^33^. A proportion of first trimester miscarriages where no chromosomal abnormalities are detected could have a Mendelian or polygenic origin^34,35^.

We previously hypothesised that many human genes contributing to prenatal lethality are likely unidentified and not captured in current disease databases due to early embryo losses and miscarriages either being unnoticed, or when they are detected, the difficulty in determining the molecular basis of this extreme phenotype. Here, we used a set of 624 genes associated with early lethality in humans curated from OMIM^6,29^. 19 % of EL disease associated genes are linked to pre and perinatal lethality. For LL genes, this percentage is 31% (**Fig 3i**). Based on our hypothesis that most genes associated to early gestation lethality in humans remain unrecognised, the set of EL in the mouse constitutes a source of candidates of interest in the field of foetal precision medicine.

#### 2.3. Predicting new EL genes in the mouse

Since the number of IMPC mouse lines that have undergone the primary viability assessment is higher than those with a secondary evaluation to identify the embryonic stage at which lethality occurs, we tried to predict additional EL genes among lethal genes without secondary viability data to have a larger pool of candidate genes. For this we used a penalised likelihood approach to fit a generalised additive model using proliferation (essentiality) scores from multiple human cell lines as predictors^22^ and subsequently used that model to make the predictions. This added an additional set of 362 predicted EL genes (out of 725 lethal genes with no secondary viability assessment) to the previous 430 EL genes assessed through embryo viability screening. Details on the model, predictive accuracy, and predictor variables can be found in Methods and **Sup Fig 5**. Out of 33 genes in our prediction set that were externally assessed as EL^36^, 29 were correctly predicted by the classifier (87.9%) (**See Sup File 2**). CRISPR knockout screens to identify those genes affecting cell survival across hundreds of genomically characterized cancer cell lines^37^ can consequently assist the identification of early gestation lethal lines in the mouse.

#### 2.4. Similarity with known BIEM genes

A gene similarity strategy was applied to 792 (assessed and predicted) EL genes based on features shared with 552 diagnostic-grade BIEM genes from PanelApp. This approach was based on the unknown gene sharing at least one of 5 attributes: p1) being a paralogue of a known BIEM gene; p2) sharing a pathway with a BIEM gene; p3) belonging to the same protein complex as a known BIEM gene; p4) interacting with another gene in the protein-protein interaction network of a known BIEM gene; and/or p5) sharing a PFAM protein family with a known BIEM gene. This gene ranking approach serves a dual purpose: 1) to identify completely novel disease genes, and 2) to bring additional proof for those genes in PanelApp that are not considered diagnostic-grade genes, i.e. ‘amber’ and ‘red’ genes. Among novel EL genes not associated with any disease in PanelApp, 53 - 60% share at least one of the five attributes with a BIEM gene. This percentage increases to 69 - 74% when the non-diagnostic-grade genes in PanelApp excluding the IEM panel are examined and to 100% for the non-diagnostic grade genes on the IEM panel (**Fig 4a**).

**Fig 4.**
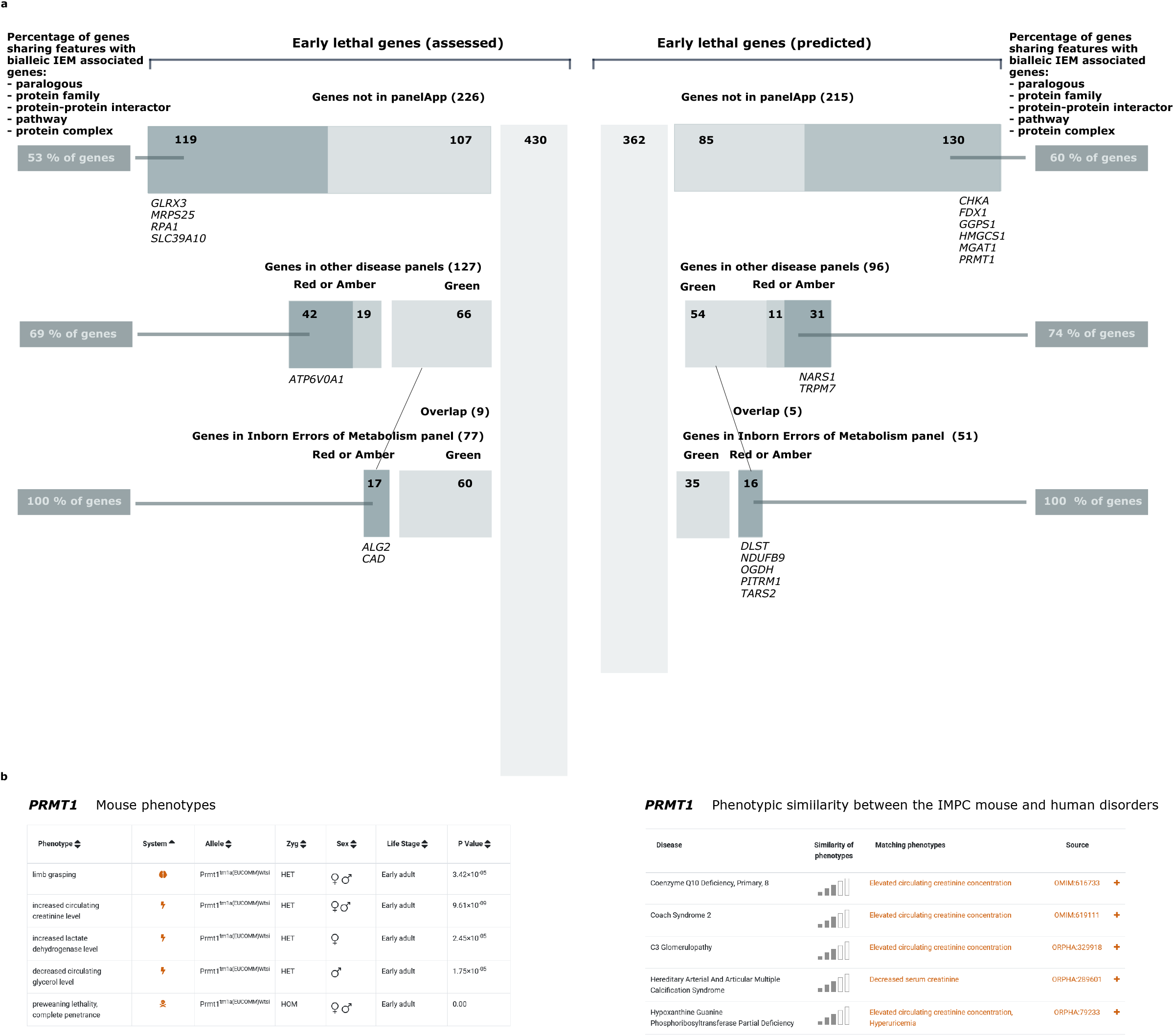
Gene similarity approach. **4a** For each set of EL genes in the mouse (assessed and predicted), the total number of genes is broken down into 3 categories based on PanelApp evidence: genes associated with inborn errors of the metabolism, Mendelian disease genes in other disease categories and non-disease genes. For genes in PanelApp panels the genes are also subdivided into those with strong evidence for the gene-disease association (green) and those with more limited evidence to date (red or amber). The percentage of genes sharing features with known BIEM genes is shown for potential novel genes not present in PanelApp as well as those with more limited evidence. For each category, those genes sharing ≥4 features with known BIEM genes are shown. **4b *PRMT1* IMPC mouse phenotypes and phenotypic similarity with human disorders**. Heterozygous knockout phenotypes include several metabolic and neurological abnormalities. When computing the similarity between the mouse and human disease phenotypes associated with known disorders, we find phenotypic overlap with several early onset conditions, including defects of the metabolism Coenzyme Q10 deficiency, primary, 8 and Hypoxanthine guanine phosphoribosyltransferase partial deficiency. EL, early gestation lethal; BIEM, biallelic inborn errors of the metabolism; IMPC, International Mouse Phenotyping Consortium.

Ten of the EL non-disease-associated genes are of particular interest as they share 4 of the 5 attributes with BIEM genes: *CHKA, FDX1, GGPS1, GLRX3, HMGCS1, MGAT1* and *SLC39A10* are paralogous and direct interactors as well as belonging to the same protein family(ies) and pathway(s) whilst *MRPS25, PRMT1*, and *RPA1* are interactors, share a protein family(ies) and pathway(s) and are also part of the same protein complex(es). The complete gene list and annotations are provided in **Sup File 3**. Four of these genes, *GGPS1, MRPS25, PRMT1*, and *RPA1* show abnormal metabolic phenotypes in the heterozygous viable mouse. *MRPS25* is a member of the human mitochondrial ribosomal protein gene family, with evidence from mouse embryos indicating compromised mitochondrial function^38^. Several other mitochondrial ribosomal small (MRPS) and large (MRPL) subunit genes are associated with different metabolic disorders, and many of the remaining MRPS genes are also potentially associated with disease^39^. Evidence of pathogenicity of homozygous missense variants in this gene has been reported^40^. In the case of *PRMT1*, encoding a member of the protein arginine N-methyltransferase (PRMT) family, additional neurological phenotypes imply a high phenotypic similarity with neonatal disorders including several defects of the metabolism as computed by PhenoDigm (**Fig 4b**). Emerging evidence supports the role of this family of enzymes in skeletal muscle and metabolic disease^41^.

Importantly, we found a significant association between sharing any of these 5 attributes with a BIEM gene and being EL (1.64 fold-increase, P value = 2.7e-06). When these attributes were considered separately, the strongest association was observed for being part of the same protein complex as a BIEM gene (13.9 fold-increase, P value = 6.5e-20). Significant results were also obtained for sharing a pathway and interacting with a BIEM gene. EL genes were less likely to be a paralogue of a BIEM gene (OR = 0.49, P value = 0.018), which can be explained by the enrichment for singletons among this set of genes (**Sup Fig 6**).

Disaggregating the set of EL genes by disease association showed that the closer to the IEM disease class, the higher the percentage of genes in that category sharing attributes with BIEM genes. Consistently, EL genes are more likely to share attributes with BIEM genes compared to non-EL genes.

#### 2.5. Undiagnosed cases of inherited metabolic disorders from the 100KGP

An alternative approach, based on patient data, was also used to identify potential metabolic disease genes among the set of EL genes in the mouse. Cases recruited under the ‘undiagnosed metabolic disorder’ and ‘mitochondrial disorders’ categories in the 100KGP were investigated for rare, segregating, and biallelic LoF or predicted pathogenic missense variants in EL genes, using the Exomiser variant prioritisation tool^11^. Observed versus expected ratios (oe) per gene were computed by comparing the number of biallelic variants observed in these patients to those observed on a set of *pseudo controls*, i.e., patients recruited under other disease categories. Predicted homozygous or compound heterozygous pathogenic variants were found in 21 EL genes (13 assessed, 8 predicted) with oe ratios > 1 and observed in ≤ 2 controls. Three involved biallelic LoF, 6 had biallelic LoF/missense, and 12 had biallelic missense variants. Five of these genes are already classified as diagnostic grade genes in the IEM panel (*COQ4, ELAC2, MRPL44, MSTO1*, and *SKIV2L*) and three others are diagnostic grade genes in different neurology and neurodevelopmental disorder gene panels (*EIF2B4, ELP1, EXOSC8*). *ALG2, NDUFA8*, and *RNASEH2A* are classified as amber or red in the IEM panel. For the cases associated with these 11 known disease genes, only those associated with *MRPL44* and *ALG2* biallelic variants have been diagnosed with these variants so far, with the others currently classified as variants of uncertain significance. For the remaining 10 genes (*AFDN, CDK12, COQ3, GINS4, GPATCH1, INTS11, KIF2C, NUFIP1, PTPMT1, RCC1*) there is no current evidence for a disease association in PanelApp or OMIM.

For two of the amber or red genes in the inborn errors of the metabolism panel, *ALG2* and *NDUFA8*, IMPC heterozygous knockout mice have neurological and metabolic phenotypes, providing additional evidence to validate this gene-disease association. In addition, *ALG2* shares 4 features with known BIEM genes: protein family (2 genes), pathway (10 genes), paralogue (1 gene), and protein-protein interaction (9 genes). Similarly, *NDUFA8* shares 3 features: protein complex (17 genes), pathways (44 genes), and protein-protein interaction (28 genes).

Four non-disease associated genes have IMPC data for null alleles with heterozygous mouse mimicking some of the clinical features observed in patients. *AFDN* and *NUFIP1* show neurological phenotypes in the mouse embryo or early adult. *COQ3* and *CDK12* also show neurological and other physiological system phenotypes shared between the undiagnosed patients and the knockout mouse. They are of particular interest as several other genes from the same family have already been associated with similar disorders, and the IMPC lines are the first reported mouse models with abnormal phenotypes observed in the early adult heterozygous knockout (**Fig 5**).

**Fig 5.**
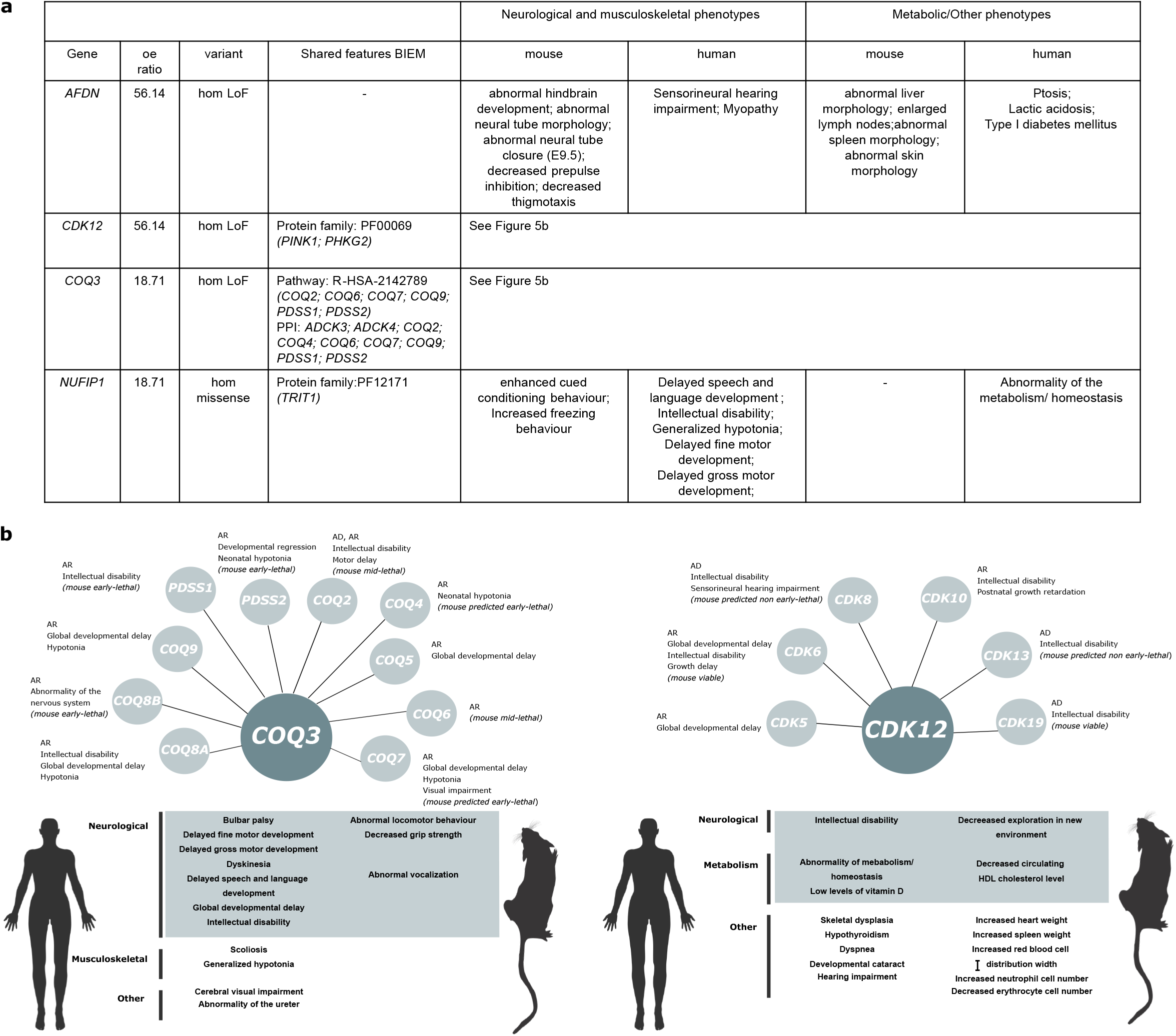
Candidate genes with biallelic inheritance involving LoF or (missense) predicted pathogenic variants in undiagnosed patients. **5a Mouse evidence.** Genes with homozygous LoF or missense variants found in patients recruited under the metabolic disorder disease category with an o/e ratio > 1, observed in a number of controls ≤ 2 and with the heterozygous knockout mouse displaying abnormal phenotypes in the relevant physiological systems, partially mimicking the phenotypes observed in patients. **5b *COQ3* and *CDK12* belong to families and pathways with several genes associated with Mendelian disorders**. The corresponding mode of inheritance and related/overlapping phenotypes for these known disease genes and evidence on viability from the IMPC are shown. Mouse phenotypes according to IMPC Data Release 15.1. LoF, loss-of-function; o/e, observed vs expected; IMPC, International Mouse Phenotyping Consortium.

*COQ3* (coenzyme Q3, methyltransferase) is one of the genes required for the biosynthesis of Coenzyme Q10, which has many vital functions. Several genes involved in this pathway are associated with Primary CoQ10 Deficiency, including *PDSS1, PDSS2, COQ2, COQ4, COQ5, COQ6, COQ7, COQ8A, COQ8B* and *COQ9*^42^. The heterozygous IMPC mouse shows several neurological / behavioural phenotypes including abnormal locomotor behaviour, abnormal vocalization, and decreased grip strength. No homozygous LoF variants have been observed for this gene according to gnomAD (pLI = 0; pRec = 0.283; DOMINO = Very likely recessive). The homozygous frameshift variant observed in the 100KGP cohort is present in gnomADv2.1.1 (p.Lys366SerfsTer2), with an allele frequency of 6.04e-04 but with no homozygous individuals for that allele. The o/e ratio in our 100KGP study cohort is 18.7, with the other two different variants found in the set of *pseudo controls* recruited under the ‘unexplained sudden death in the young’ and ‘ultra-rare undescribed monogenic disorders’.

C*DK12* (cyclin dependent kinase 12) is one of the cyclin-dependent kinases with a key role in molecular processes relevant during development. Several other protein kinases are involved in developmental disorders: *CDK5, CDK6, CDK8, CDK10, CDK13* and *CDK19*^43^. The phenotypic abnormalities observed in heterozygous IMPC mice include cardiac, hematopoietic, metabolic (decreased circulating HDL cholesterol level), and neurological features (decreased exploration in new environment) (**Fig 5b**). The homozygous splice acceptor variant (c.1047-2A>G) is present in gnomADv2.1.1, with an allele frequency of 4.06e-4 and one homozygote observed in the South Asian population. This gene is in fact predicted to be highly intolerant to heterozygous LoF variation (pLI = 1; pRec = 0; DOMINO = Very likely dominant). The o/e ratio computed with biallelic variants in our GEL study cohort for this gene is 56.14 with no variants meeting the criteria described found in controls.

A note of caution is needed when interpreting the impact of these two LoF variants due to their position on the transcript, as indicated by gnomAD. Where available, gene expression across development data for the four genes confirm similar trajectories across mouse and human for brain and development and intolerance to LoF variation.

The approach described here is based on the premise that biallelic LoF in a gene leads to early embryonic lethality in mice but that biallelic LoF or missense variants in humans lead to recessively inherited metabolic disorders with related phenotypes in humans (**Fig 3b, 3c**). In fact, for the four candidate genes highlighted in our study, it is the heterozygous mouse model which is mimicking the phenotypes observed in patients carrying biallelic mutations.

This somehow counterintuitive observation has been reported for other IEM disorders^44,45^. When exploring early lethality annotations in humans, up to 39% of the known biallelic IEM genes have records of lethality before or shortly after birth, showing that a considerable proportion of these conditions in humans are life threatening, leading to early death if untreated. Consistent with this observation, two of the genes in the same pathway or gene family of our candidate genes (*COQ9, PDSS2)* have been associated with early lethality in humans^46,47^.

## Discussion

Many predicted LoF variants identified in Mendelian disease sequencing studies are found in genes not previously associated with disease, making assessment of pathogenicity particularly challenging. High throughput mouse standardised phenotyping screens including viability assessment contribute to acquiring new knowledge about orthologues of such genes with limited functional data^48,49^. By also exploring correlations between abnormal phenotype(s) in the knockout mouse and disease features in the human orthologues, we are able to identify novel candidates for Mendelian conditions.

Previously we developed a successful framework to prioritise gene candidates for neurodevelopmental disorders using mouse phenotyping data, with two of the top 9 candidate genes, *VPS4A* and *SPTBN1*, already having been recently validated. In both cases, a causal link has been found between heterozygous, predominantly *de novo* mutations and distinctive developmental syndromes^16–18^. Here we present another example of how the IMPC data resource can be combined with other sources of evidence to develop a tailored approach for disease-gene discovery and variant prioritisation to assist the diagnosis of inherited metabolic disorders.

The requirement of a gene for the survival of an organism, i.e. gene essentiality, can be disaggregated into more granular categories/WoL according to the embryonic period during which lethality occurs. In the present study, we show that these categories, correlate with different gene features, including gene expression across development and intolerance to LoF variation. Additionally, the distribution of singleton and duplicated genes across these WoL support hypotheses about genetic compensation. EL genes are more likely to be singletons, and when paralogues exist they tend to have originated earlier, suggesting more time to evolve new functions^26,27^.

By looking at different features of human disease genes across the windows, two observations stand out. First, the set of lethal genes in the mouse is enriched for Mendelian disease genes^15^, but the proportion of genes associated with disease is not consistent across WoL and this enrichment is mainly driven by LL genes. The lower proportion of disease genes among the EL compared to LL genes was previously reported when comparing cellular lethal with developmental lethal genes^16^, as well as other categorisations of essential genes^29,50^. The contribution of lethal genes to human disease, including embryonic lethality as a genuine Mendelian phenotype^51^, is probably understated, and the lower proportion of disease genes among the EL is potentially due to an underrepresentation of genes leading to embryonic lethal human phenotypes in current disease databases. Second, we identified a strong association between EL genes and inherited metabolic disorders. This includes genes that are needed to maintain the metabolic machinery required to provide energy and basic components for cell survival. Most of the EL lines die prior to implantation or gastrulation, and differentiation into disease associated tissues occurs at a later stage. This could explain why non-metabolic disease categories are underrepresented among the set of EL genes.

Building on this finding, we focused on the EL genes and gathered additional information on similarity with known disease genes associated with biallelic forms of inborn errors of metabolism (BIEM). It is already known that members of paralogous gene families where one gene is associated to human disease are more likely to be associated with Mendelian disorders themselves^52^. Similarly, disease associated variants are enriched at sites conserved among paralogues^53,54^. We used these and other observations to identify the EL genes showing most similarity to existing BIEM genes and, hence, most likely to be novel BIEM disease genes.

Inherited metabolic disorders comprise a large group of ∼1,450 disorders in which the primary alteration of a biochemical pathway leads to a set of biochemical, clinical, and/or pathophysiological features^55^. The majority manifest in new-borns, show predominantly neurological manifestations and can lead to sudden premature death^56^. By investigating patients recruited under this disease category from the 100KGP and looking at EL genes in the mouse for evidence of enrichment of biallelic LoF or predicted pathogenic missense variants, we were able to identify a set of candidate genes where the heterozygous knockout mouse mimicked some neurological and/or metabolic phenotypes observed in patients.

Two of the genes identified through our analysis, *COQ3* and *CDK12*, belong to pathways and extended gene families of genes which are associated with similar disorders, which strongly supports their involvement in the disease process. Further functional characterisation of these and other predicted pathogenic variants, together with the identification of additional probands with biallelic variants segregating with similar phenotypes are still needed to establish a causal link, to confirm that the candidate LoF variants result in the lack of protein product and/ or have a discernible clinical phenotypic effect.

Comparing lethality outcomes between mouse and human presents several limitations. Monoallelic mutations required for early development (dominant lethals) are missing from our set of mouse embryonic lethal knockouts since they would not result in lines, introducing a bias towards recessive lethal genes. Prenatal lethality is the most severe phenotypic manifestation found in monogenic forms of disease. Disrupted gene function may lead to embryonic lethality in humans at very early stages, which makes it difficult to recognise, since many of these early pregnancy losses go undetected^57^. Most metabolic disorders represent a spectrum of phenotypes. According to OMIM clinical records, more than a third of BIEM genes are associated with lethality before or soon after birth. This may help explain the differences between mouse and human, with the homozygous knockout showing early embryonic lethality and the heterozygous mouse mimicking patient phenotypes.

Underestimation of prenatal lethality as the most severe phenotype among a broader range of severe clinical manifestations reported in humans seems a plausible hypothesis for some discrepancies in lethality observed between the two species. Similarly, while in the mouse knockouts the observed phenotype is most likely due to the loss of protein function, other types of mutation may lead to different molecular mechanisms and thus different phenotypic outcomes. True loss of protein function in these genes may be early embryonic lethal in humans whereas postnatal phenotypes could be caused by hypomorphic variants leading to partial LoF^58,59^. Other explanations include potential mechanisms of compensation through other genes in the pathway in humans or differences in essentiality between the two species. Even when gene essentiality does not perfectly correlate, the mouse models provide knowledge on the molecular functions and biological processes^60^. Given the number of genes associated with lethality in the mouse (35% of the knockout lines are classified as lethal or subviable according to IMPC primary viability screening)^15,16^, monogenic factors could explain a proportion of the high and often understated level of occurrence of miscarriages in human^34^. Therefore, the set of lethal genes in the mouse constitutes an invaluable resource to identify relevant genes in humans, including those in which LoF variation may lead to pregnancy loss and other severe phenotypes with an early manifestation^29,57^.

In summary, the embryonic stage at which lethality occurs in the mouse can be used to inform human disease. Integration of multi-species datasets and the extended use of standardised phenotypes is key to building novel Mendelian gene discovery approaches^3,61^. This, coupled with the availability of large-scale sequencing programs that allow for bespoke computational and statistical analysis for variant prioritisation constitutes a powerful instrument for increasing the molecular diagnostic rate^21^. Every time a new gene is linked to a specific Mendelian condition, numerous series of undiagnosed patients could be revisited world-wide, which may translate into new genetic diagnoses. The findings of this study and previous research will expedite the development of new gene identification strategies tailored to specific types of disorders. Additionally, the set of genes essential for embryonic development in the mouse may constitute an additional source of evidence for diagnosis of lethal foetal disorders^29,62,63^. Whether this is the only observable outcome or the most extreme phenotype within a wider range of clinical features observed in patients, it will be crucial to catalogue these genes.

Ultimately, our work complements other strategies for identification of novel genes underlying Mendelian conditions. It also highlights the power of cross-species phenotype analysis by integrating model organism resources with data from large scale sequencing programs.

## Methods

### Data sources

#### IMPC mouse data

Mouse primary and secondary viability data were obtained from the IMPC resource^64^. Primary viability data: http://ftp.ebi.ac.uk/pub/databases/impc/all-data-releases/release-15.0/results/viability.csv.gz (DR15) [Downloaded 28.09.21]

Phenotype annotations: https://www.mousephenotype.org/ (DR15.1) [Accessed 02.11.21] Embryonic viability data: Detailed information on the primary and secondary viability pipelines, including definitions, procedures, and protocols can be found at https://www.mousephenotype.org/impress/index. These include: Viability Primary Screen, Viability E9.5 Secondary Screen, Viability E12.5 Secondary Screen, Viability E14.5-E15.5 Secondary Screen, Viability E18.5 Secondary Screen, Homozygote Viability at Weaning Screen.

#### Entire set of human protein coding genes with the corresponding mouse orthologs

One to one human orthologues with the corresponding HGNC identifiers were obtained from the HUGO Gene Nomenclature Committee (HGNC) resource^65^: http://ftp.ebi.ac.uk/pub/databases/genenames/hgnc/tsv/locus_groups/protein-coding_gene.txt [Downloaded 28.09.21]. All the following gene features used in this study correspond to human gene annotations. Gene symbols, Ensemble and Uniprot ids were converted into HGNC unique identifiers. Where there was any ambiguity about gene id mapping, the annotation was discarded.

#### Human cell proliferation scores

CRISPR knockout screens from the Achilles pipeline (release 21Q3) for 902 cell lines and the corresponding cell line information were obtained from the DepMap portal^22^: https://depmap.org/portal/download/all/ (Achilles_gene_effect_CERES.csv) [Downloaded 28.09.21]. Gene effect scores are direct estimates of the effect of a gene knockout on viability. Thus, a more negative CERES score indicates more depletion in the cell line. Average scores per gene were computed. In order to establish a binary threshold to classify genes as cellular essential and non-essential, previous data on cell essentiality, based on 11 cell lines from 3 different studies was used to compute F1 scores derived from confusion matrices generated when considering different CERES mean scores and the classification from these 3 studies, and a mean score cut-off of −0.40, −0.45, and −0.55 was found to maximise the F1 scores across the different datasets, similar to the −0.45 threshold estimated with information from 485 cell lines^16,64^.

#### Gene expression across development

Human gene expression (RPKM) across development for brain, cerebellum, heart, kidney, liver, ovary and testis was obtained from Cardoso-Moreira et al.^23^ https://apps.kaessmannlab.org/evodevoapp/ [Downloaded 10.08.21] Data on comparison of temporal trajectories between human genes and their orthologs in mouse for brain and cerebellum was obtained from Cardoso-Moreira et al.^25^.

#### Intolerance to variation scores

gnomaAD v2.1.1 constraint metrics^66^ (pLI and pRec) and DOMINO scores^67^: https://gnomad.broadinstitute.org/downloads#v2constraint ; https://wwwfbm.unil.ch/domino/ [Downloaded 10.08.21]

#### Gene duplicates

Information of paralogues of human genes was obtained from Ensembl Biomart (Ensembl Genes 104)^68^ https://www.ensembl.org/biomart/martview/ Only protein coding paralogues with HGNC ids and % amino acid identity >=20% were considered [Downloaded 10.08.21]

#### Protein-protein interactions

Human protein network data (scored links between proteins) was obtained from STRING^69^ https://stringdb.org/cgi/downloadãsessionId=%24input%3E%7BsessionId%7D&species_text=Homo+sapiens [Downloaded 13.08.21]

#### Pathways

Lowest level pathways were obtained from Reactome^70^ https://reactome.org/download/current/UniProt2Reactome.txt and https://reactome.org/download/current/ReactomePathways.txt [Downloaded 10.08.21]

#### Protein families

PFAM protein families^71^ were obtained through Ensembl biomart (Ensembl Genes 104) https://www.ensembl.org/biomart/martview/ [Downloaded 10.08.21]

#### Protein complex

Corum protein complex information^72^ was accessed at: https://mips.helmholtzmuenchen.de/corum/#download [Downloaded 13.08.21]

### Disease features

#### Mendelian disease genes, disease category and mode of inheritance

Diagnostic grade ‘green’ genes with sufficient evidence for disease association and their corresponding modes of inheritance were obtained from Genomics England PanelApp, a publicly-available knowledge base containing panels related to human disorders^31^. A total number of 313 gene panels (excluding additional findings) were investigated. Information on allelic requirement and level of evidence of disease causation was retrieved for our analysis. Genes from 186 gene panels containing level 2 disease category information (21 categories) were used for the analysis based on disease classification. https://PanelApp.genomicsengland.co.uk/panels/ [Downloaded 10.08.21]

#### HPO phenotypes

Phenotypes were obtained from the HPO (genes to phenotypes)^73^ and mapped to the top level of the ontology, broadly corresponding to the physiological system affected. Co-occurrence with the most frequent systems affected (neurological and musculoskeletal) were computed for early lethal genes (EL) versus non early lethal genes (NEL). https://hpo.jax.org/app/download/annotation; https://raw.githubusercontent.com/obophenotype/human-phenotype-ontology/master/hp.obo [Downloaded 23.08.21, HPO notes: format-version: 1.2 data-version: hp/releases/2021-08-02]

#### Prenatal and perinatal genes in humans

A set of 624 genes associated to prenatal and perinatal lethality based on OMIM records were used for the analysis^6,29^.

### Prediction of early lethal genes

Several genes have undergone the IMPC primary viability assessment, but the embryonic stage at which lethality occurs has not yet been investigated. To increase the pool of potential candidate early lethal genes, we built a classifier using human cell proliferation scores from 902 lines as predictor variables. For that we used the R implementation of Generalized Additive Model Selection, *gamsel*. The training set consisted of 893 genes, 430 early-lethal (EL) and 463 non-early lethal (NEL). Imputation of missing values was performed via nuclear-norm regularization implemented in the *softImpute* R package. Cross validation (5-fold) was used to assess the performance of the model, with ROC-AUCs ranging from 0.860 to 0.903. The accuracy ranged from 79.9 to 86.0% of instances correctly classified as EL and NEL. Predictions for a total number of 725 lethal genes with no secondary viability assessment were made. Only 24 predictor variables (non-zero effects) were selected in the final model. A probability threshold of 0.41 maximized the F1 score. Using this model, 362 genes were predicted as EL and the remaining 363 as NEL (**Sup Fig 5**). A set of 33 genes externally assessed as EL^36^ was used as additional validation (**Sup File 3**).

### Gene similarity approach

Similarity with known genes associated to biallelic forms of inherited metabolic disorders (biallelic inborn error of metabolism green genes from PanelApp, BIEM) was assessed according to 5 attributes (5ps): p1) being a paralogue of a known BIEM gene according to Ensembl genes 104 and a threshold of % amino acid identity of 20%^68^; p2) sharing a Reactome pathway (lowest level) with a BIEM gene^70^; p3) belonging to the same Corum protein complex of a BIEM gene^72^; p4) being a direct interactors within the protein-protein interaction network (high confidence cut-off 0.7) with a BIEM gene according to STRING^69^; p5) sharing a PFAM protein family with a BIEM gene^71^.The number of different features shared was computed for every early lethal gene - assessed and predicted.

### Investigation of cases from the 100KGP

To investigate the occurrence and enrichment of homozygous LoF variants in cases from the 1000KGP among our set of EL genes in the mouse we searched for variants in those genes in 35,422 families, 631 of which were recruited under the categories of interest (‘undiagnosed metabolic disorders’ and ‘mitochondrial disorders’). One important caveat is that these are not healthy population controls, and we cannot rule out that patients recruited under other categories show similar metabolic phenotypes, which means that these ratios can be an underestimation. The number of observed homozygous LoF and missense variants prioritised by Exomiser based on variant scores^11^ were compared between cases and *pseudo* controls to compute observed versus expected ratios.

### Software

R software^74^ including the following packages were used for data integration and analysis: *tidyverse*^75^, *matrixStats*^76^, *epitools*^77^; data visualization: *waffle*^78^, *ggridges*^79^, *alluvial*^80^, *cowplo*t^81^, *upSetR*^82^ ; ontologies: *ontologyIndex*^83^; modelling and prediction: *softImpute*^84^, *gamsel*^85^, *pROC*^86^. Odds Ratios were calculated by unconditional maximum likelihood estimation (Wald) and confidence intervals (CI) using the normal approximation, with the corresponding adjusted P-values (BH) for the test of independence using the *oddsratio* function. To test for significant differences in proportions in the different WoL, *prop*.*test* and *pairwise*.*prop*.*test* functions were applied. In the case of continuous variables, to test for significant differences between the three WoL we used the Kruskal-Wallis and Dunn’s test implementations: *kruskal*.*test* and *dunnTest* functions.

## Data availability

The supplementary files supporting the findings of this study are made publicly available through Zenodo: https://doi.org/10.5281/zenodo.5796622.

Viability reports and additional files containing mouse embryo and adult phenotypes are available through the IMPC web portal (https://www.mousephenotype.org/) and the IMPC FTP repository (http://ftp.ebi.ac.uk/pub/databases/impc/).

## Ethical approval

The IMPC Consortium collects data from international member institutes who collect phenotyping data guided by their own ethical review panels, licenses, and accrediting bodies that reflect the national and/or geo-political constructs in which they operate (Institutional Animal Care and Usage Committee, Baylor College of Medicine; Animal Welfare and Ethical Review Body (AWERB), MRC Harwell; Animal Care Committee (ACC) of The Centre for Phenogenomics; The Jackson Laboratory Institutional Animal Care and Use Committee (IACUC); UC Davis Institutional Animal Care and Use Committee (IACUC)).

All the information regarding animal ethics approval of mouse production, breeding and phenotyping, including study design, experimental procedures, housing and husbandry and sample size can be found in the following links:

https://www.mousephenotype.org/about-impc/animal-welfare/

https://www.mousephenotype.org/about-impc/animal-welfare/arrive-guidelines/

All efforts were made to minimize suffering by considerate housing and husbandry. All phenotyping procedures were examined for potential refinements that were disseminated throughout the Consortium. Animal welfare was assessed routinely for all mice involved.

All patient data used from the 100,000 Genomes Project were accessed through the research environment provided by Genomics England and conforming to their procedures. All participants in the 100KGP have provided written consent to provide access to their anonymised clinical and genomic data for research purposes.

## Data Availability

The supplementary files supporting the findings of this study are made publicly available through Zenodo: https://doi.org/10.5281/zenodo.5796622.
Viability reports and additional files containing mouse embryo and adult phenotypes are available through the IMPC web portal (https://www.mousephenotype.org/) and the IMPC FTP repository (http://ftp.ebi.ac.uk/pub/databases/impc/).

## Acknowledgments

This work was supported by NIH grant U54 HG006370 (P.C, C.H.W., V.M-F., J.M., H.M.M., H.P, A-M.M., D.S). Other National Institute of Health grants include R01 HD083311 (J.M), UM1 HG006348 (M.E.D, C.H., J.D.H, L.T, S.W), UM1 OD023221 (C.M. K.C.K.L., L.L), UM1 OD023221-09S1 (C.M.), UM1 OD0023222 (S.A.M.), U42 OD011174 and 5UM1 HG006348-10 (L.T, S.W., J.C., R.B.S., M.S., J.H.). Additional support was provided by the Medical Research Council, Strategic Award A410-53658 (L.T, S.W., J.C., R.B.S., M.S., J.H.). This research was made possible through access to the data and findings generated by the 100,000 Genomes Project (http://www.genomicsengland.co.uk). The 100,000 Genomes Project is managed by Genomics England Limited (a wholly owned company of the Department of Health and Social Care). The 100,000 Genomes Project is funded by the National Institute for Health Research and NHS England. The Wellcome Trust, Cancer Research UK and the Medical Research Council have also funded research infrastructure. The 100,000 Genomes Project uses data provided by patients and collected by the National Health Service as part of their care and support and we are grateful to both for making this available.

## Contributions

P.C., D.S. contributed to conceptualization, data analysis, presentation and interpretation of the results and writing the manuscript. C.H.W. contributed to data analysis, interpretation, reviewing and editing the manuscript. J.M. contributed to data generation, interpretation, reviewing and editing the manuscript. M.E.D, V.M.-F., I.B.V.d.V., J.D.H contributed to data interpretation, reviewing and editing the manuscript. L.M.J.N, A.M.F supervised the research generating the embryos, viability and windows of lethality data, reviewed and edited the manuscript. C.-W.H. contributed to data analysis and presentation. C.M. contributed to reviewing and editing the manuscript. S.A.M., K.C.K.L, L.L. contributed to data collection, interpretation, editing and reviewing the manuscript. L.T., S.W contributed to conceptualization, data generation and data interpretation. A.K.C., V.L., R.G, D.Q performed the research generating embryos, viability and windows of lethality data. J.C, R.B.-S., M.S, J.H. contributed to data generation and interpretation. J.M., H.H.M. contributed to data analysis and software development. M.E.D, C.M., J.D.H, K.C.K.L, R.E.B., J.K.W., A-M.M., H.P., D.S. are PIs of the key programmes who contributed to the management and execution of the work. The additional IMPC members all contributed to data acquisition and data handling.

## International Mouse Phenotyping Consortium

John R. Seavitt^5^, Angelina Gaspero^5^, Uche Akoma^4^, Audrey Christiansen^4^, Sowmya Kalaga^4^, Lance C. Keith^4^, Melissa L. McElwee^4^, Leeyean Wong^4^, Tara Rasmussen^4^, Uma Ramamurthy^4,14,15^, Kiran Rajaya^14^, Panitee Charoenrattanaruk^14^, Qing Fan-Lan^6^, Lauri G. Lintott^6^, Ozge Danisment^6^, Patricia Castellanos-Penton^6^, Daniel Archer^12^, Sara Johnson^12^, Zsombor Szoke-Kovacs^12^, Kevin A. Peterson^11^, Leslie O. Goodwin^11^, Ian C. Welsh^11^, Kristina J. Palmer^11^, Alana Luzzio^11^, Cynthia Carpenter^11^, Coleen Kane^11^, Jack Marcucci^11^, Matthew McKay^11^, Crystal Burke^11^, Audrie Seluke^11^, Rachel Urban^11^

^14^ Office of Research Information Technology, Baylor College of Medicine, Houston, TX, USA

^15^ Department of Pediatrics, Baylor College of Medicine, Houston, TX, USA

## Genomics England Research Consortium

Ambrose, J. C.^16^ ; Arumugam, P.^16^ ; Bevers, R.^16^ ; Bleda, M.^16^ ; Boardman-Pretty, F.1^1,16^ ; Boustred, C. R.^16^ ; Brittain, H.^16^ ; Caulfield, M. J.^1,16^ ; Chan, G. C.^16^ ; Fowler, T.^16^ ; Giess A.^16^ ; Hamblin, A.^16^ ; Henderson, S.^1,16^ ; Hubbard, T. J. P.^16^ ; Jackson, R.^16^ ; Jones, L. J.^1,16^ ; Kasperaviciute, D.^1,16^ ; Kayikci, M.^16^ ; Kousathanas, A.^16^ ; Lahnstein, L.^16^ ; Leigh, S. E. A.^16^ ; Leong, I. U. S.^16^ ; Lopez, F. J.^16^ ; Maleady-Crowe, F.^16^ ; McEntagart, M.^16^ ; Minneci F.^16^ ; Moutsianas, L.^1,16^ ; Mueller, M.^1,16^ ; Murugaesu, N.^16^ ; Need, A. C.^1,16^ ; O’Donovan P.^16^ ; Odhams, C. A.^16^ ; Patch, C.^1,16^ ; Perez-Gil, D.^16^ ; Pereira, M. B.^16^ ; Pullinger, J.^16^ ; Rahim, T.^16^ ; Rendon, A.^16^ ; Rogers, T.^16^ ; Savage, K.^16^ ; Sawant, K.^16^ ; Scott, R. H.^16^ ; Siddiq, A.^16^ ; Sieghart, A.^16^ ; Smith, S. C.^16^ ; Sosinsky, A.^1,16^ ; Stuckey, A.^16^ ; Tanguy M.^16^ ; Taylor Tavares, A. L.^16^ ; Thomas, E. R. A.^1,16^ ; Thompson, S. R.^16^ ; Tucci, A.^1,16^ ; Welland, M. J.^16^ ; Williams, E.^16^ ; Witkowska, K.^1,16^ ; Wood, S. M.^1,16^

^16^ Genomics England, London, UK

