## Supplementary Data for "Mendelian gene identification through mouse embryo viability screening"

**Cacheiro et al.**

### Content

**Supplementary Figures 1-6**

**Supplementary Tables 1-3**

**Sup Fig 1 WoL and cell essentiality scores**. **1a CERES depletion scores for blood lineage human cell lines across WoL**. A more negative scores indicates more depletion of the gene in the cell line, i.e. more essential. Triangles represent median values of gene expression per WoL. **1b CERES depletion scores for ovary lineage human cell lines across WoL.** A more negative scores indicates more depletion of the gene in the cell line, i.e. more essential. Triangles represent median values of gene expression per WoL. **1c Binary categorization of CEREs depletion scores** . Genes are categorised as essential vs non-essential based on a threshold to maximize F1 score using a binary classification from previous dataset. **1d WoL and cellular essential genes**. Percentage of EL, ML and LL genes considered cellular essential when alternatives -0.40 and -0.55 thresholds are considered. WoL, windows of lethality; EL, early gestation lethal; ML, mid gestation lethal; LL, late gestation lethal.

**Sup Fig 2 WoL and organ development trajectories**. Percentage of genes in each WoL with the same organ development trajectory between mouse and human (brain and cerebellum). The grey dashed line shows the baseline %, corresponding to the percentage of genes showing the same trajectory for the entire set of genes with data available. WoL, windows of lethality.

**Sup Fig 3 WoL and additional gene features**. **3a WoL and mode of inheritance**. Predicted mode of inheritance according to DOMINO scores, that assess the likelihood for a gene to harbour dominant change. **3b WoL, paralogues and cellular essentiality.** Singleton genes are more likely to be cellular essential compared to duplicate genes in the same window. WoL, windows of lethality.

**Sup Fig 4 WoL and additional disease categories**. Frequency of genes for the remaining disease categories across WoL. The grey dashed line shows the baseline %, corresponding to the percentage of genes in each disease with respect to the total number of PanelApp “green” genes (3,384). WoL, windows of lethality.

**Sup Fig 5 Prediction of early lethal genes**. A penalised likelihood approach was used to fit a generalised additive model using proliferation (essentiality) scores from multiple cell lines as predictors and subsequently used that model to make the predictions. The effect of each variable is estimated to be either zero, linear, or a low-complexity curve). **5a ROC-AUC**. 5-fold CV ROC-AUC and accuracy estimated a balanced set of 895 genes (430 EL, 465 NEL). **5b Predictor variables**. Human cancer cell lines with DepMap CERES scores with non-zero coefficients. Only 24 cell lines out of 902 were selected in the final model. CV, Cross-validation; ROC-AUC, Area Under the Receiver Operating Characteristic Curve; EL, early gestation lethal; NEL, non-early gestation lethal (mid gestation lethal and late gestation lethal).

Sup Fig 6 **Enrichment analysis of genes sharing attributes with a BIEM gene among the EL category**. Odds Ratios with 95% confidence intervals and BH adjusted P values for the five attributes investigated. EL genes were compared to ANEL genes (mid gestation lethal, late gestation lethal, subviable and viable genes). EL, early lethal genes; ANEL, all non-early gestation lethal genes.

Sup Table 1 **Gene features.** Test for differences among WoL. P values for comparison across the three groups and post hoc pairwise comparisons.

Sup Table 2 **Disease features.** Test for differences among WoL. P values for comparison across the three groups and post hoc pairwise comparisons.

Sup Table 3 **HPO phenotypes Odds Ratios**. Odds Ratio with 95% confidence intervals and BH adjusted P values for EL genes compared to NEL genes (mid gestation lethal + late gestation lethal). No significant differences were found for any of the top level HPO phenotypes, corresponding to physiological systems. EL, early gestation lethal; HPO, human phenotype ontology.

### Sup Fig 1

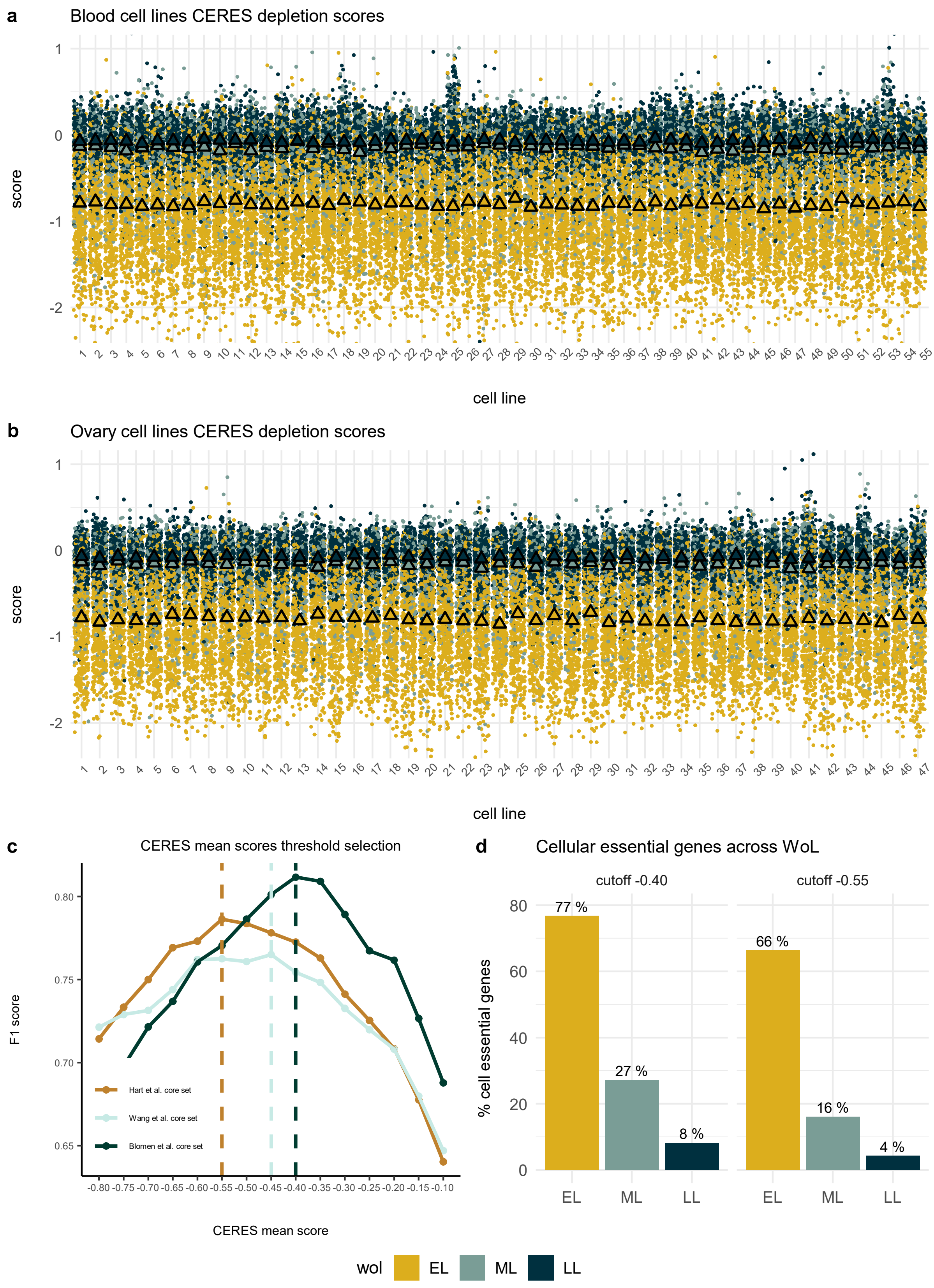
a
Blood cell lines CERES depletion scores
score
cell line
b
Ovary cell lines CERES depletion scores
score
cell line
c
CERES mean scores threshold selection
F1 score
CERES mean score
Hart et al. core set
Wang et al. core set
Blomen et al. core set
d
Cellular essential genes across WoL
cutoff -0.40
cutoff -0.55
% cell essential genes
77 %
27 %
8 %
66 %
16 %
4 %
EL
ML
LL
wol
EL
ML
LL

### Sup Fig 2

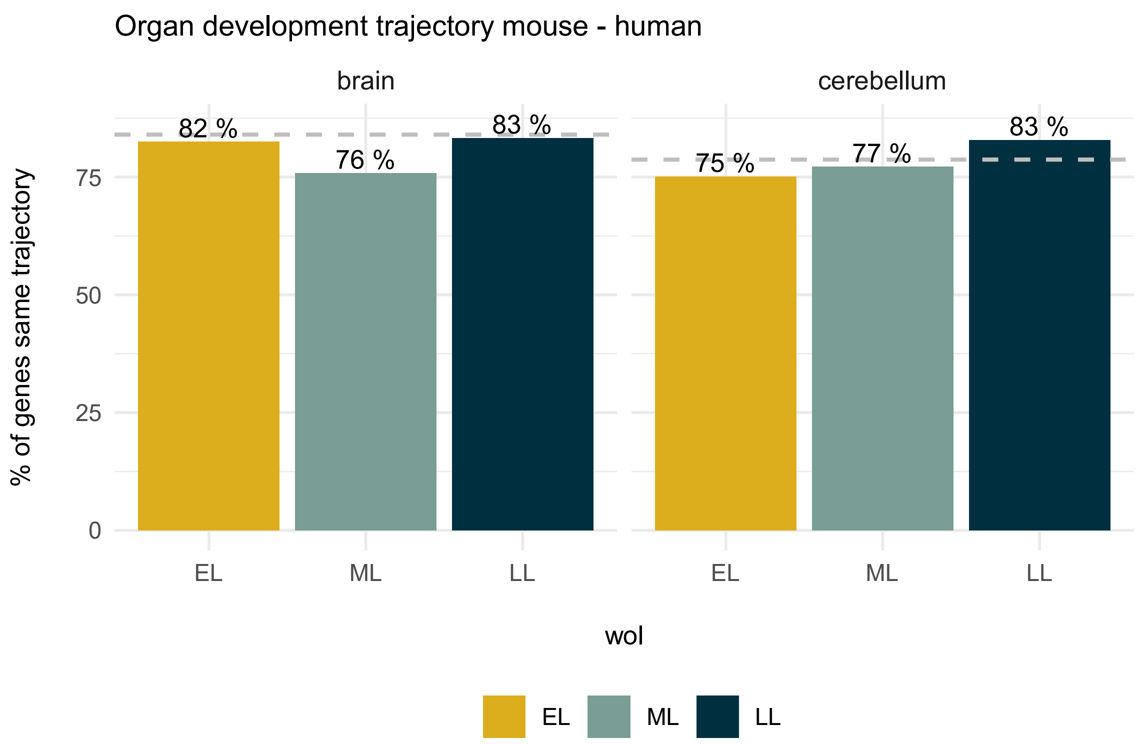
Organ development trajectory mouse - human
brain
cerebellum
82 %
76 %
83 %
75 %
77 %
83 %
% of genes same trajectory
0
25
50
75
EL
ML
LL
EL
ML
LL
wol
EL
ML
LL

### Sup Fig 3

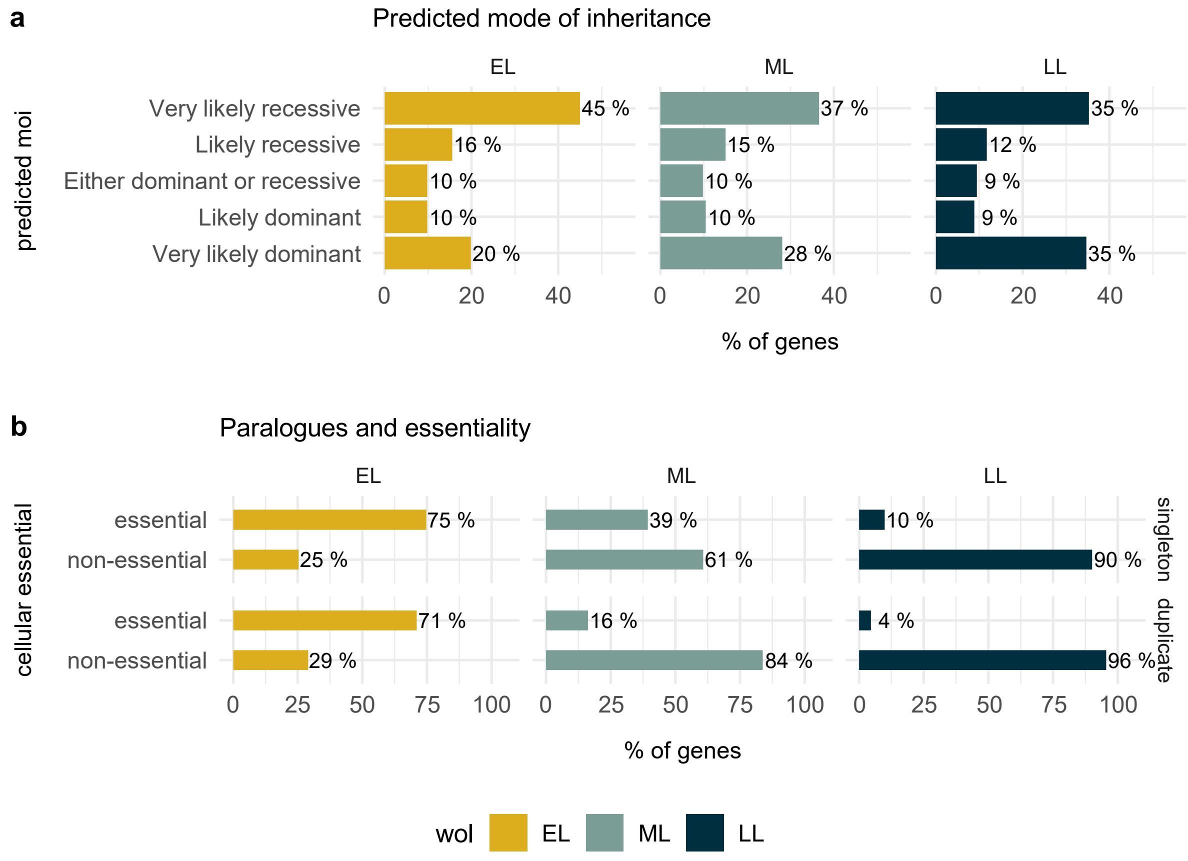
a
Predicted mode of inheritance
EL
ML
LL
predicted moi
Very likely recessive
Likely recessive
Either dominant or recessive
Likely dominant
Very likely dominant
45 %
16 %
10 %
10 %
20 %
37 %
15 %
10 %
10 %
28 %
35 %
12 %
9 %
9 %
35 %
0 20 40
% of genes
b
Paralogues and essentiality
EL
ML
LL
cellular essential
essential
non-essential
essential
non-essential
singleton
duplicate
75 %
25 %
71 %
29 %
39 %
61 %
16 %
84 %
10 %
90 %
4 %
96 %
0 25 50 75 100
% of genes
wol
EL
ML
LL

### Sup Fig 4

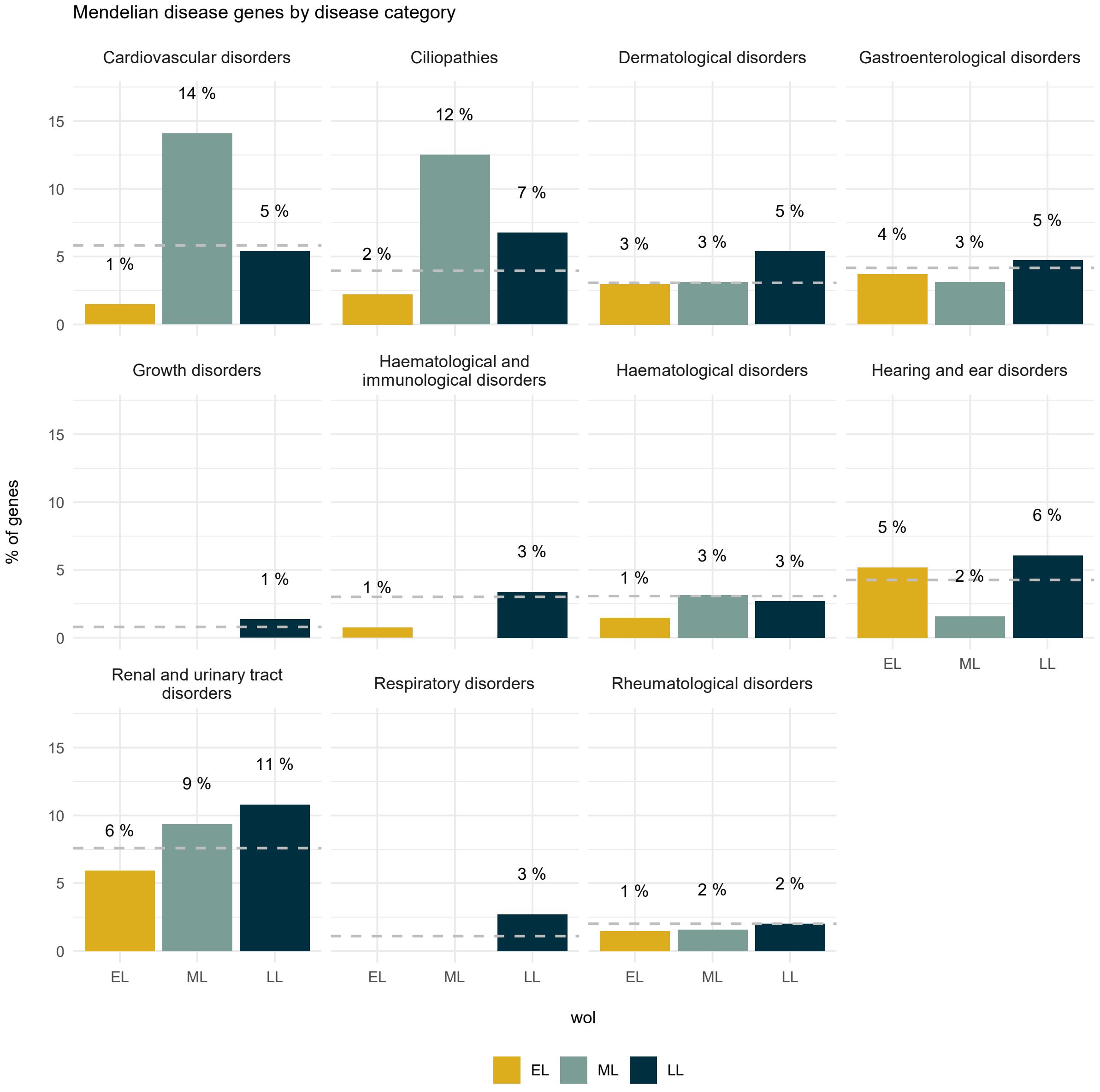

Mendelian disease genes by disease category
Cardiovascular disorders
Ciliopathies
Dermatological disorders
Gastroenterological disorders
Growth disorders
Haematological and immunological disorders
Haematological disorders
Hearing and ear disorders
Renal and urinary tract disorders
Respiratory disorders
Rheumatological disorders
14 %
1 %
5 %
12 %
2 %
7 %
3 %
3 %
5 %
4 %
3 %
5 %
1 %
1 %
3 %
1 %
3 %
3 %
5 %
2 %
6 %
6 %
9 %
11 %
3 %
1 %
2 %
2 %
% of genes
0
5
10
15
EL
ML
LL
wol
EL
ML
LL

### Sup Fig 5

a

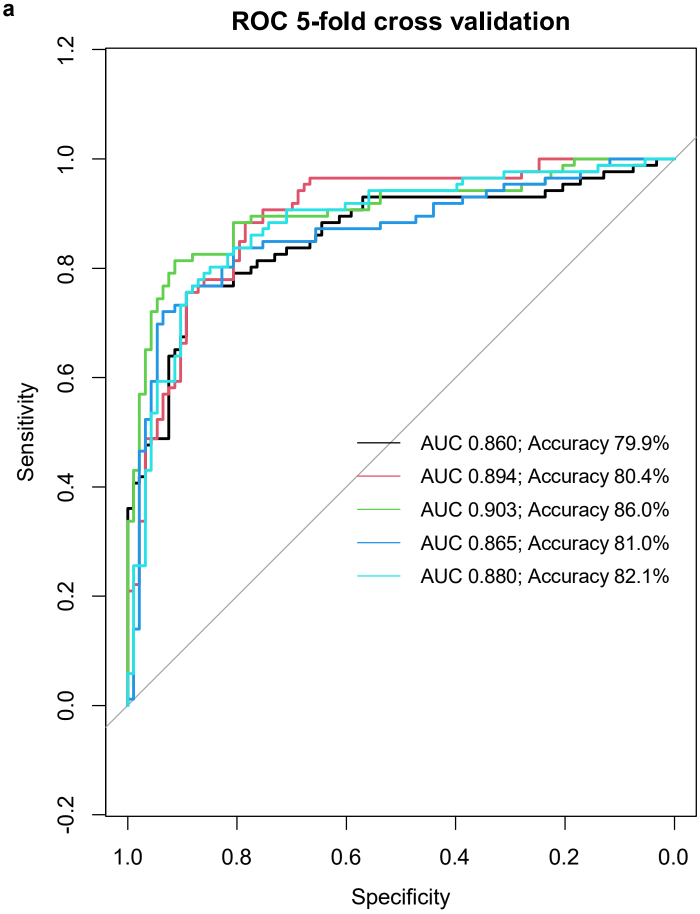


b

| | DepMap_ID | lineage |
|---|---|---|
| *1* | ACH-000045 | blood |
| *2* | ACH-000141 | bile_duct |
| *3* | ACH-000257 | lung |
| *4* | ACH-000272 | kidney |
| *5* | ACH-000313 | kidney |
| *6* | ACH-000318 | esophagus |
| *7* | ACH-000459 | kidney |
| *8* | ACH-000537 | liver |
| *9* | ACH-000573 | breast |
| *10* | ACH-000618 | upper_aerodigestive |
| *11* | ACH-000677 | lung |
| *12* | ACH-000760 | central_nervous_system |
| *13* | ACH-000851 | lung |
| *14* | ACH-000969 | colorectal |
| *15* | ACH-001041 | lung |
| *16* | ACH-001061 | colorectal |
| *17* | ACH-001163 | kidney |
| *18* | ACH-001232 | central_nervous_system |
| *19* | ACH-001307 | thyroid |
| *20* | ACH-001518 | uterus |
| *21* | ACH-001628 | ovary |
| *22* | ACH-001688 | kidney |
| *23* | ACH-001737 | blood |
| *24* | ACH-001786 | colorectal |

### Sup Fig 6

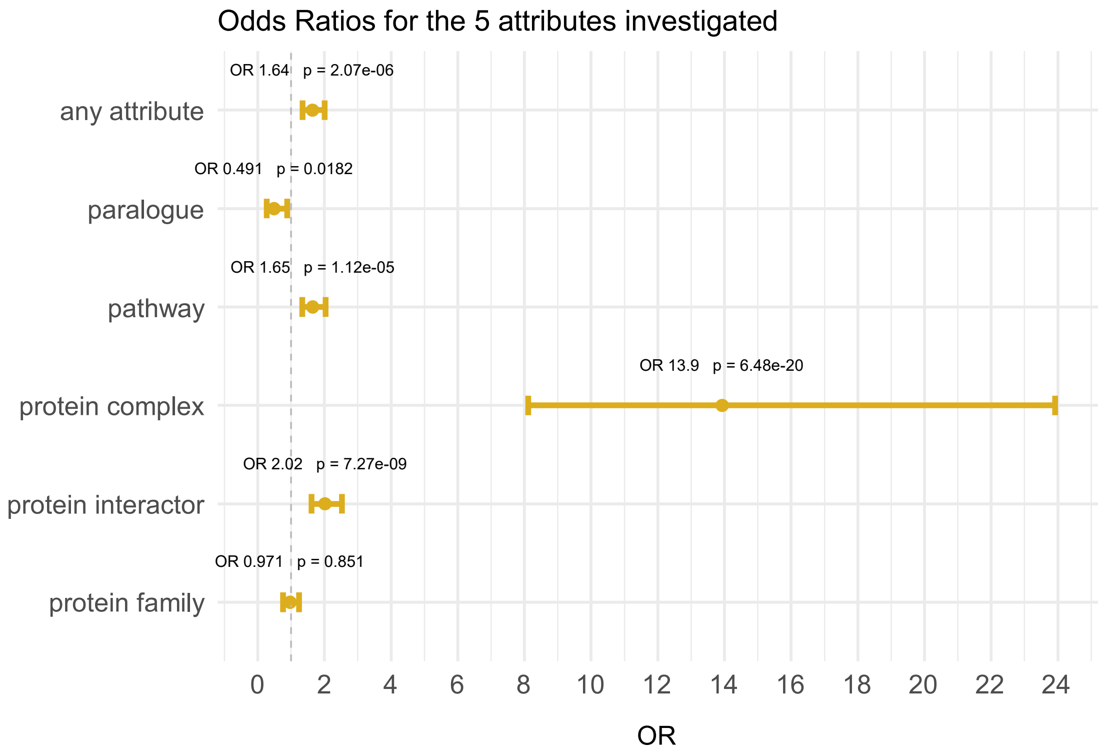
Odds Ratios for the 5 attributes investigated
OR 1.64 p = 2.07e-06
any attribute
OR 0.491 p = 0.0182
paralogue
OR 1.65 p = 1.12e-05
pathway
OR 13.9 p = 6.48e-20
protein complex
OR 2.02 p = 7.27e-09
protein interactor
OR 0.971 p = 0.851
protein family
0 2 4 6 8 10 12 14 16 18 20 22 24
OR

### Sup Table 1

| Feature | Figure | Early-Mid-Late | Early-Mid | Mid-Late | Early-Late |
|---|---|---|---|---|---|
| CNS proliferation scores | 2a | < 2.2e-16 | < 2.2e-16 | < 2.2e-16 | < 2.2e-16 |
| Blood proliferation scores | S1a | < 2.2e-16 | < 2.2e-16 | < 2.2e-16 | < 2.2e-16 |
| Ovary proliferation scores | S1b | < 2.2e-16 | < 2.2e-16 | < 2.2e-16 | < 2.2e-16 |
| Cellular essential | 2c | < 2.2e-16 | < 2.2e-16 | 7.4e-09 | < 2e-16 |
| Gene expression brain 4wpc | 2d | < 2.2e-16 | 7.0e-05 | 7.3e-03 | 1.6e-17 |
| Gene expression brain 10wpc | 2d | 4.4e-12 | 1.4e-05 | 2.7e-01 | 5.3e-12 |
| Gene expression brain 20wpc | 2d | 1.7e-07 | 1.1e-03 | 3.4e-01 | 8.5e-08 |
| Gene expression brain infant | 2d | 1.1e-06 | 7.2e-03 | 1.8e-01 | 2.7e-07 |
| Gene expression brain senior | 2d | 5.7e-07 | 6.5e-03 | 1.6e-01 | 1.4e-07 |
| pRec | 2e | 0.047 | 0.038 | 0.599 | 0.038 |
| pLI | 2f | 0.011 | 0.007 | 0.347 | 0.029 |
| Singletons | 2g | < 2.2e-16 | 1.1e-04 | 2.6e-03 | < 2.2e-16 |
| Opisthokonta | 2h | 1.4e-09 | 4.7e-04 | 0.204 | 5.2e-10 |
| Bilateria | 2h | 1.0e-05 | 0.012 | 0.272 | 2.8e-06 |
| Paralogues | 2i | 9.4e-05 | 5.1e-03 | 6.6e-01 | 3.1e-05 |

Uncorrected P values are shown. Alpha threshold for significance was set at 0.0005 given the number of hypothesis tested.

### Sup Table 2

| Feature | Figure | Early-Mid-Late | Early-Mid | Mid-Late | Early-Late |
|---|---|---|---|---|---|
| Mendelian genes | 3a | 3.1e-05 | 0.033 | 0.223 | 9.1e-06 |
| MoI | 3b | 1.3e-05 | 0.008 | 0.418 | 5.2e-06 |
| Neurology | 3c | 0.513 | 1.000 | 0.500 | 0.370 |
| Metabolic | 3c | 2.7e-06 | 0.025 | 0.152 | 1.1e-06 |
| Skeletal | 3c | 0.046 | 0.026 | 0.436 | 0.116 |
| Ophthalmological | 3c | - | - | - | - |
| Dysmorphic | 3c | 0.510 | 0.600 | 1.000 | 0.340 |
| Endocrine | 3c | 0.210 | 0.370 | 1.000 | 0.130 |
| Cardiovascular | S3a | - | - | - | - |
| Ciliopathies | S3a | - | - | - | - |
| Dermatological | S3a | - | - | - | - |
| Gastroenterological | S3a | - | - | - | - |
| Growth | S3a | - | - | - | - |
| Haematological | S3a | - | - | - | - |
| Hearing | S3a | - | - | - | - |
| Renal | S3a | 0.335 | 0.550 | 0.940 | 0.210 |
| Respiratory | S3a | - | - | - | - |
| Rheumatological | S3a | - | - | - | - |

Uncorrected P values are shown. Alpha threshold for significance was set at 0.0005 given the number of hypothesis tested. Number of observations was too small in some groups given the low number of disease associated genes for certain categories.

### Sup Table 3

| | hpo | or | or_lower | or_upper | pvalue | pvalue.adjust |
|---|---|---|---|---|---|---|
| 1 | Abnormality of the nervous system | 1.3616595 | 0.29143759 | 6.364895 | 0.7220459 | 0.8443480 |
| 2 | Abnormality of head or neck | 1.6943917 | 0.75052980 | 3.861061 | 0.2199826 | 0.5241434 |
| 3 | Abnormality of metabolism/homeostasis | 0.3720294 | 0.07545236 | 1.335207 | 0.1459925 | 0.5241434 |
| 4 | Growth abnormality | 1.9768317 | 0.82829991 | 4.807708 | 0.1282873 | 0.5241434 |
| 5 | Abnormality of the integument | 1.6613629 | 0.76075839 | 3.681376 | 0.2382470 | 0.5241434 |
| 6 | Abnormality of the genitourinary system | 0.5393277 | 0.23687958 | 1.196682 | 0.1622182 | 0.5241434 |
| 7 | Abnormality of the respiratory system | 0.7849718 | 0.35784085 | 1.712733 | 0.5594431 | 0.8205165 |
| 8 | Abnormality of limbs | 0.9355834 | 0.42739397 | 2.049569 | 1.0000000 | 1.0000000 |
| 9 | Abnormality of the digestive system | 0.7909173 | 0.30821405 | 1.954409 | 0.6534132 | 0.8443480 |
| 10 | Abnormality of the endocrine system | 0.8577810 | 0.37098518 | 1.996348 | 0.8322241 | 0.8718538 |
| 11 | Abnormality of the cardiovascular system | 1.2123952 | 0.53485815 | 2.740426 | 0.6829483 | 0.8443480 |
| 12 | Abnormality of blood and blood-forming tissues | 1.3789226 | 0.62175084 | 3.111461 | 0.5462543 | 0.8205165 |
| 13 | Abnormality of the immune system | 0.5901894 | 0.25899617 | 1.332392 | 0.2199826 | 0.5241434 |
| 14 | Abnormality of the musculoskeletal system | 0.6603267 | 0.12539804 | 2.749587 | 0.7292096 | 0.8443480 |
| 15 | Abnormality of prenatal development or birth | 0.6902324 | 0.27136615 | 1.750949 | 0.4847161 | 0.8205165 |
